# Modeling the impact of the Omicron infection wave in Germany

**DOI:** 10.1101/2022.07.07.22277391

**Authors:** Benjamin F. Maier, Angelique Burdinski, Marc Wiedermann, Annika H. Rose, Matthias an der Heiden, Ole Wichmann, Thomas Harder, Frank Schlosser, Dirk Brockmann

## Abstract

**BACKGROUND:** In November 2021, the first case of SARS-CoV-2 “variant of concern” (VOC) B.1.1.529 (“Omicron”) was reported in Germany, alongside global reports of reduced vaccine efficacy against infections with this variant. The potential threat posed by the rapid spread of this variant in Germany remained, at the time, elusive.

**METHODS:** We developed a variant-dependent population-averaged susceptible-exposed-infected-recovered (SEIR) infectious disease model. The model was calibrated on the observed fixation dynamics of the Omicron variant in December 2021, and allowed us to estimate potential courses of upcoming infection waves in Germany, focusing on the corresponding burden on intensive care units (ICUs) and the efficacy of contact reduction strategies.

**RESULTS:** A maximum median incidence of approximately 300 000 (50% PI in 1000: [181,454], 95% PI in 1000: [55,804]) reported cases per day was expected with the median peak occurring in the mid of February 2022, reaching a cumulative Omicron case count of 16.5 million (50% PI in mio: [11.4, 21.3], 95% PI in mio: [4.1, 27.9]) until Apr 1, 2022. These figures were in line with the actual Omicron waves that were subsequently observed in Germany with respective peaks occurring in mid February (peak: 191k daily new cases) and mid March (peak: 230k daily new cases), cumulatively infecting 14.8 million individuals during the study period. The model peak incidence was observed to be highly sensitive to variations in the assumed generation time and decreased with shorter generation time. Low contact reductions were expected to lead to containment. Early, strict, and short contact reductions could have led to a strong “rebound” effect with high incidences after the end of the respective non-pharmaceutical interventions. Higher vaccine uptake would have led to a lower outbreak size. To ensure that ICU occupancy remained below maximum capacity, a relative risk of requiring ICU care of 10%–20% was necessary (after infection with Omicron vs. infection with Delta).

**CONCLUSIONS:** We expected a large cumulative number of infections with the VOC Omicron in Germany with ICU occupancy likely remaining below capacity nevertheless, even without additional non-pharmaceutical interventions. Our estimates were in line with the retrospectively observed waves. The results presented here informed legislation in Germany. The methodology developed in this study might be used to estimate the impact of future waves of COVID-19 or other infectious diseases.

## 1. INTRODUCTION

During the COVID-19 pandemic, infectious disease modeling proved to be an essential tool to estimate the impact of upcoming waves of the disease under different scenario assumptions regarding pathogen properties as well as pharmaceutical and non-pharmaceutical interventions [1– 3]. Knowledge about the order of magnitude of an upcoming crisis can help to inform legislation regarding interventions to prevent public health systems and critical infrastructure from overburdening and collapse [3–5]. With continuously emerging SARS-CoV-2 variants of concern that can have relatively heterogeneous properties like virulence and severity of disease, model-based analyses continue to provide valuable insights on upcoming challenges and potential mitigation strategies [6].

Prior to the “variant of concern” (VOC) B.1.1.529 (“Omicron”) becoming the dominant strain of SARS-CoV-2 in Germany during the COVID-19 pandemic, we therefore modeled possible trajectories of the upcoming infection wave in 2022 in Germany, taking into account a variety of parameter estimates calibrated on the growth behavior of Omicron cases and cases of the VOC B.1.617.2 (“Delta”) still prevalent in December 2021. Despite sustained high vaccine effectiveness against severe courses of the disease [7], reduced efficacy against infection was suspected to lead to higher growth rates and large outbreaks and, therefore, a potentially high burden on the healthcare system and critical infrastructure [8]. In order to estimate this potential impact the spread of Omicron could have in Germany, and how potential non-pharmaceutical and pharmaceutical (i.e. vaccination) interventions might affect the trajectory of its spread, we devised and analyzed a parsimonious infectious disease model in this study. Our analysis was based, at the time, on the (partially limited) available empirically data regarding the properties of the VOCs Omicron and Delta as well vaccine efficacies and expectations of the booster distribution campaign. Here, we publish and reflect on our results that were originally published as a technical report in German on Feb 3, 2022 [9].

Our model analysis predicted, at the time, peak incidences and outbreak sizes that were in line with the later observed waves. We found that the model peak height strongly depended on variations in the assumed generation time and decreased with shorter generation time, which highlights the importance of reliable empirical values of this epidemiological parameter. Regarding the efficacy of contact-reducing non-pharmaceutical interventions, even lower contact reductions were expected to lead to containment, early, strict, and short contact reductions, however could have had an adverse effect after the respective lifting of restrictions at a later time, due to a waning of population-averaged vaccine efficacy. Based on our results, we estimated that the relative risk (RR) of requiring intensive care for an infection with Omicron compared to an infection with Delta must assume values of the order of 10%–20% to prevent recurrence of extreme intensive care unit (ICU) burden, which was later found to be the case [10–12]. A hypothetically higher number of first-time immunizations (in our example, 15 million additional first-time immunizations) would have, in turn, greatly reduced the risk of large waves and maximally burdened ICUs, as well. Our results informed legislation in Germany, and are, retrospectively, in good agreement with the actual course of the pandemic in Germany during the first quarter of 2022. We argue that the simple, yet effective, methodology used herein will be useful to estimate the impact of forthcoming waves of COVID-19 or other infectious diseases.

## 2. METHODS

The model followed a population-averaged susceptible-exposed-infected-recovered (SEIR) dynamic, in which no explicit distinction was made between vaccinated and unvaccinated individuals (see full methodology, Eqs. (1)–(4)). Instead, the impact of vaccines was modeled using population-averaged time-dependent vaccine efficacies (Figs. 8–9). Mathematically, this approach yields the same growth rate as models that explicitly differentiate between vaccinated and unvaccinated persons, but, in doing so, it systematically overestimates the magnitude of large outbreaks by approximately 10% in the worst case (see full methodology, Fig. 14). The advantage of this simplified approach was that the model could be quickly adjusted and analyzed, as well approximated analytically, allowing for an adaptive response to changes in the data.

Assumed time-dependent courses of vaccine efficacies are shown in Fig. 2. Since mRNA vaccines account for the absolute majority of vaccine doses in Germany, only vaccine efficacies of BioNTech and Moderna vaccines were considered in the following. We also assumed that immune evasion of the Omicron variant corresponded to preliminary data from Denmark (hereinafter referred to as “low VE”, low vaccine efficacy) [13]. For a secondary analysis, we instead chose time courses of efficacy against infection that were functionally similar to time courses of efficacy against symptomatic disease, mostly observed in the UK [7]. This corresponded to a scenario with lower immune evasion and stronger effect of booster vaccination, i.e., an optimistic scenario (hereafter referred to as “high VE”).

We further assumed that the booster vaccination campaign reached (i) 80% of individuals (referred to as “medium reach”) or (ii) all individuals who received full vaccination protection in 2021 (referred to as “high reach”). The number of newly completed vaccination series (“2 doses”) was ignored for the main analyses, but, for an illustrative analysis, the percentage was momentarily raised to approx. 90% (by artificially increasing the number of initially vaccinated persons by 15 million) to show a hypothetical course of a scenario with a high initial immunization rate.

We varied the mean latency (2 days for Delta, as well as (i) 2 days for Omicron and (ii) 1 day for Omicron) and mean infectious period (upper bound: 3 days, lower bound: 2 days), i.e., simulated scenarios for generation times of 5 days, 4 days, and 3 days. We defined the latency period as the mean duration between infection and onset of infectiousness.

The model was calibrated up to January 1, 2022 (contact modulation and VEs). To this end, the temporal contact modulation was inferred by mapping the model incidence to the observed incidence for the respective assumptions. To emulate contact behavior similar to the observed one, stochastic simulations were performed to generate contact modulation curves that had the same statistical properties as the contact modulation observed in December 2021. A vanishing variance was assumed for mean curves. Calibration of the model using the reported incidence of Delta cases showed increasing uncertainty as the proportion of Omicron cases grew, thus rendering the calibration increasingly unstable after January 1, 2022. After this date, only the daily vaccination data up to January 22 were updated and used in the results presented.

High reported infection rates can lead to behavioral changes and contact reductions [14]. For illustrative purposes, we assumed a contact reduction of −20% relative to the original courses, as well as higher contact reductions for complementary analyses.

In the United Kingdom and the United States, low values of relative risk (RR) of severe courses from infections with Omicron versus Delta were observed [10, 11, 15]. Thus, for each scenario, we determined the range of maximal possible RRs of requiring intensive care in order to keep the ICU burden at most at the level of the burden in the previous wave (“Delta wave”). For the analyses shown in the main section, we assumed values of RR of hospitalization to be RR = 0.35 and intensive care RR = 0.15 to approximate the observed trajectories of hospitalization incidence and ICU occupancy observed in early January 2022.

The results discussed here are with regard to the originally spreading Omicron sublineage (BA.1), ignoring the influence of the sublineage BA.2, which did not spread substantially before early 2022.

## 3. RESULTS

The purpose of our analyses was to provide, at the time, order-of-magnitude estimates of central epidemiological observables and scenario comparisons with regard to variation in parameters that determined the course of the Omicron infection wave.

Combining plausible assumptions and stochastic extrapolations of the contact modulation, we expected a maximum median of 300 000 new cases per day, associated with a wide uncertainty of 180 000 or 450 000 cases per day (50% prediction interval (PI)) and 55 000 or 800 000 per day (95% PI) (see Fig. 1 and Table I). Overall, a median outbreak size (cumulative number of reported Omicron cases) of 16.5 million was expected by April 1, 2022 (50% PI: [11.4, 21.3], 95% PI: [4.1, 27.9]). This figure was expected to be an overestimation at the time, as the reported cases might have been artificially reduced by changes in test prioritization or exhaustion of reporting logistics capacities, and the outbreak size is systematically overestimated due to modeling choices. Note that the model was calibrated up to January 1, 2022. Retrospectively, approximately 14.8 million Omicron infections have been reported up to April 1, 2022 [16, 17], which is approximately 10% below our median expectation, hence rather accurate considering we expected our model to overestimate the peak size by a maximum of 10%. Similarly, the observed incidence peaks in mid February (peak: 191k daily new cases) and mid March (peak: 230k daily new cases) [16], as well as the time series of new hospitalizations and cases in ICU care, respectively, lied within expectation.

**FIG. 1.**
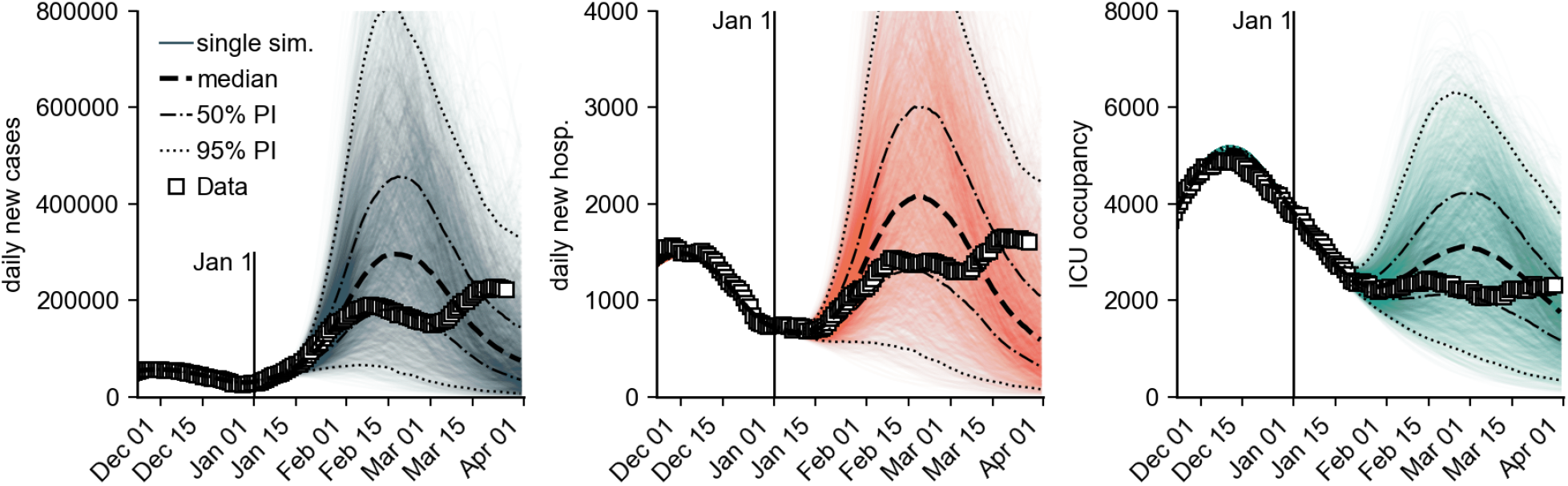
Model simulation results for the number of new cases per day, new hospitalizations per day and ICU occupancy for a combination of different model assumptions (see Methods, Sec. 2). Iterated here were “medium reach” of the booster campaign (80% of those initially immunized receive a booster vaccination), “low VE” and “high VE” of the booster (vaccine efficacy, see Methods), different generation times (5 days, 4 days (Omicron latency: 2 days), 4 days (Omicron latency: 2 days), 4 days (Omicron latency: 1 day), and 3 days), as well as “no further contact reduction” and “−20% contact reduction during the period from January 31 to March 15”. We defined the latency period as the mean duration between infection and onset of infectiousness. For each scenario combination, 180 simulations were performed, each with an individual stochastic contact modulation curve (see Methods Sec. 2)). Shown are (i) individual simulation results (colored opaque lines), (ii) the median across all model runs (black dashed line) with 50% and 95% prediction intervals, and (iii) the observed data (square data points). The relative risk of hospitalization by Omicron versus Delta was assumed to be *RR* = 0.35 and intensive care *RR* = 0.15. The model was calibrated until January 1 2022, so simulations differ from that date onward.

**FIG. 2.**
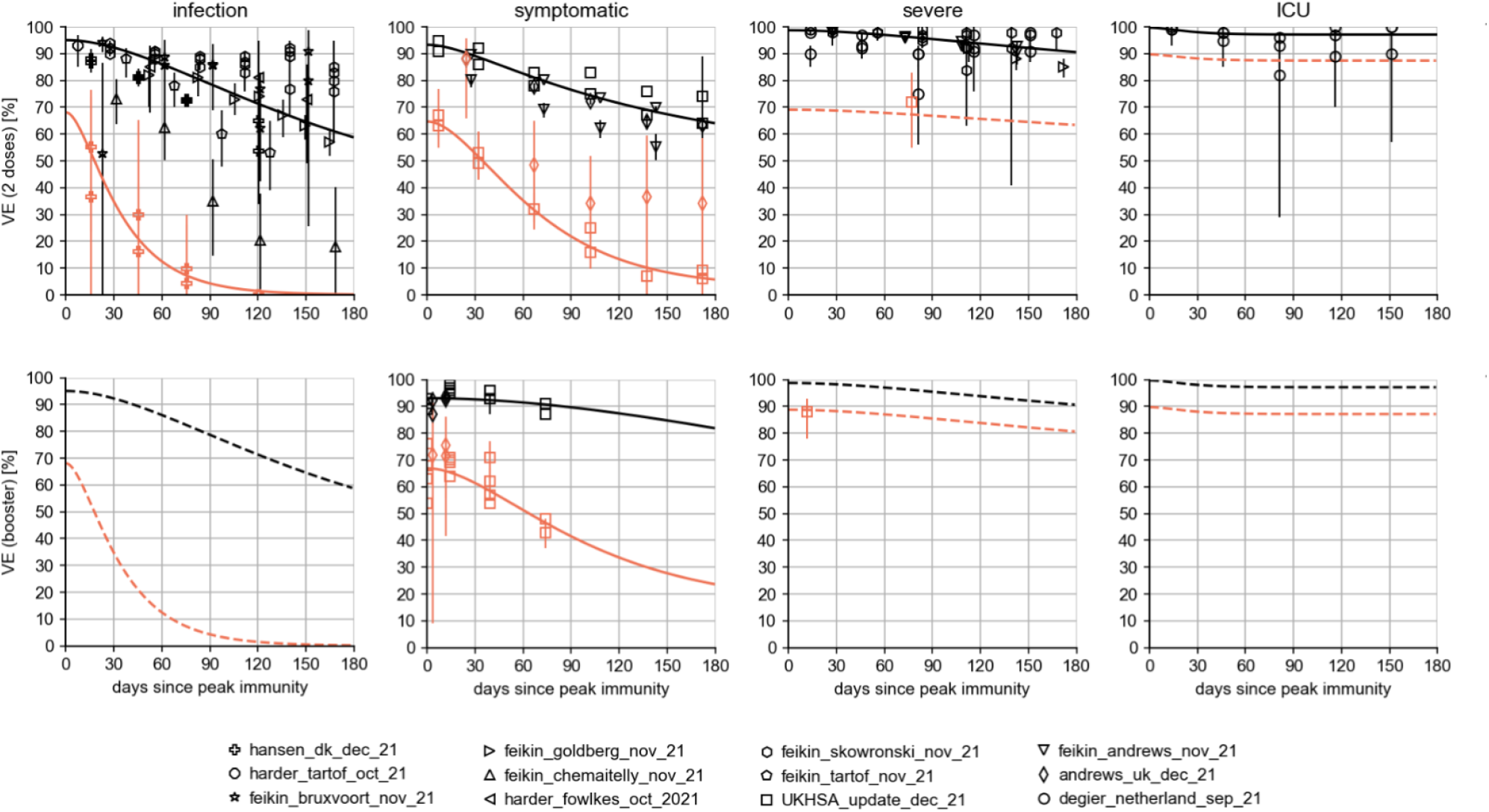
Vaccine efficacies for Omicron (orange) and Delta (black) following immunization. The top row shows the efficacies after basic immunization, the bottom row (where data were available) shows the corresponding values after booster immunization. Solid lines are the results of numerical fits; dashed lines indicate assumptions made. Data were compiled from a total of twelve studies (see list below figure and full methodology Sec. 5). For the “low VE” scenario, it was assumed that the respective VEs against *infection* after the booster dose were equal to the effect of the vaccines after receiving two doses (pessimistic). For the “high VE” scenario, we instead assumed that any VEs against infection were functionally equal to the time courses of VEs against *symptomatic disease* (optimistic).

**TABLE I.** Model prediction of the peak height of the different observables (median and prediction intervals at the date of maximum median), see Fig.1.

|  | Median [in 1000] | 50% PI | 95% PI |
| --- | --- | --- | --- |
| New cases per day | 296.2 | [180.9, 453.6] | [55.1, 803.6] |
| New hospitalizations per day | 2.1 | [1.3, 3.0] | [0.5, 4.9] |
| ICU occupancy | 3.1 | [2.1, 4.2] | [0.9, 6.3] |

Regarding the temporal evolution of the outbreak, several of the simulated outbreaks had two peaks as was then later observed in the actual outbreak, the latter peak likely being caused by the spread of the BA.2 sublineage of Omicron, which is associated with a higher base transmissibility [18]. However, most of the simulated outbreak, as well as the time series of the scenario median only shows a single peak. This scenario median time series first overestimated the daily number of new infections (in February) and subsequently underestimated them (in March).

The results were sensitive to variations in the assumed generation time (see Fig. 3). Small generation times caused larger growth rates with constant transmissibility of a variant (see Section 5 5.3). This meant that rapid increases in case numbers must be attributed to a higher basic reproduction number for longer generation times, which, in turn, caused larger model outbreaks than pathogens with shorter generation times but the same growth rate. Model results of different generation times and assumed VE (vaccine efficacy) of the booster vaccine reflected, at the time, the observed data similarly well, such that the analysis presented did not allow for conclusions about the actual contribution of the booster vaccination to the incidence. Retrospectively, the “medium reach” and “high VE” scenario shows good agreement with observed data (see Sec. 5 5.11).

**FIG. 3.**
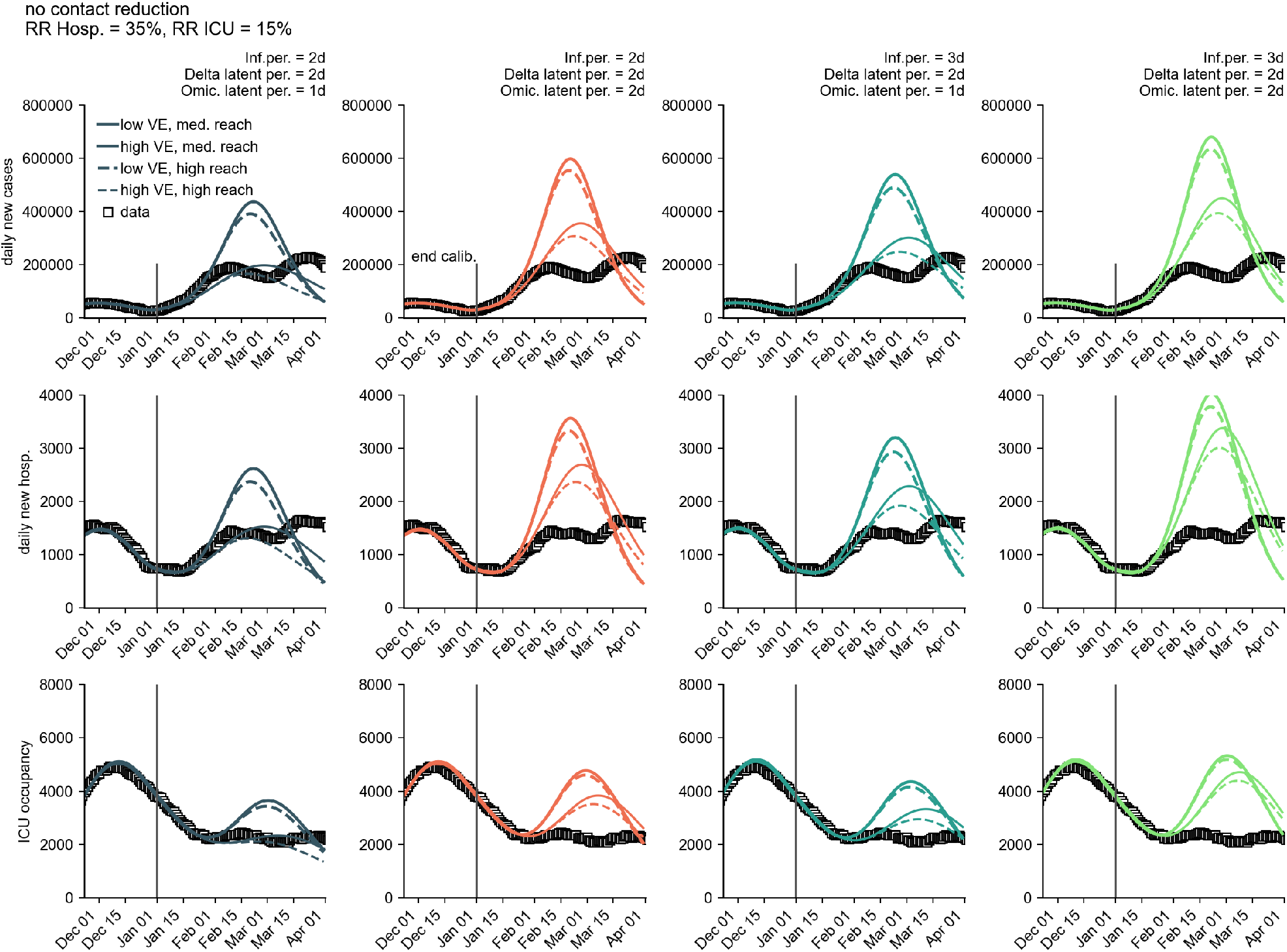
Influence of generation time on model results for the number of new cases per day (first row), new hospitalizations per day (second row) and ICU occupancy (third row) for a combination of different plausible model assumptions. Iterated here were “medium reach” and “high reach” of the booster campaign, “low VE” and “high VE” of the booster vaccination, as well as different generation times of Omicron (5 days, 4 days (Omicron latency: 2 days), 4 days (Omicron latency: 1 day) and 3 days). We defined the latency period as the mean duration between infection and onset of infectiousness. It was further assumed that no additional contact reduction occurs. For each scenario combination, a simulation was performed with an average course of the contact behavior (stochastic simulation with zero variance). Furthermore, the relative risk of hospitalization by Omicron versus Delta was assumed to be *RR* = 0.35 and intensive care *RR* = 0.15.

The model also showed a high sensitivity to the assumed booster VE against infection. The booster vaccine reach is less conclusive (within the range of 80%–100% of initially immunized individuals).

Variations in contact behavior can have a significant impact on the results, as well. Slight reductions in contact, such as those brought about by autonomous changes in the behavior of the population [14], led to substantial reductions in outbreak size in the model (see Figs. 4-5). Potentially, an additional wave could have been expected after the end of the model integration phase. This wave should have, however, been smaller due to the basic immunity in the population achieved by the first wave. This effect is illustrated by a weak, short contact reduction (−20% from January 31 to February 15), which would have had a “breaking” effect on the wave and leads to a flattening of the epidemic curve over a longer period of time. However, early, strict, yet short contact restrictions could have led to a “rebound” effect (see Fig. 4) due to a lack of population-wide immunity to infection. Such a non-pharmaceutical intervention would have caused larger outbreak sizes since the population-wide effect of the booster vaccination against infection would have already diminished by then (see Fig. 8).

**FIG. 4.**
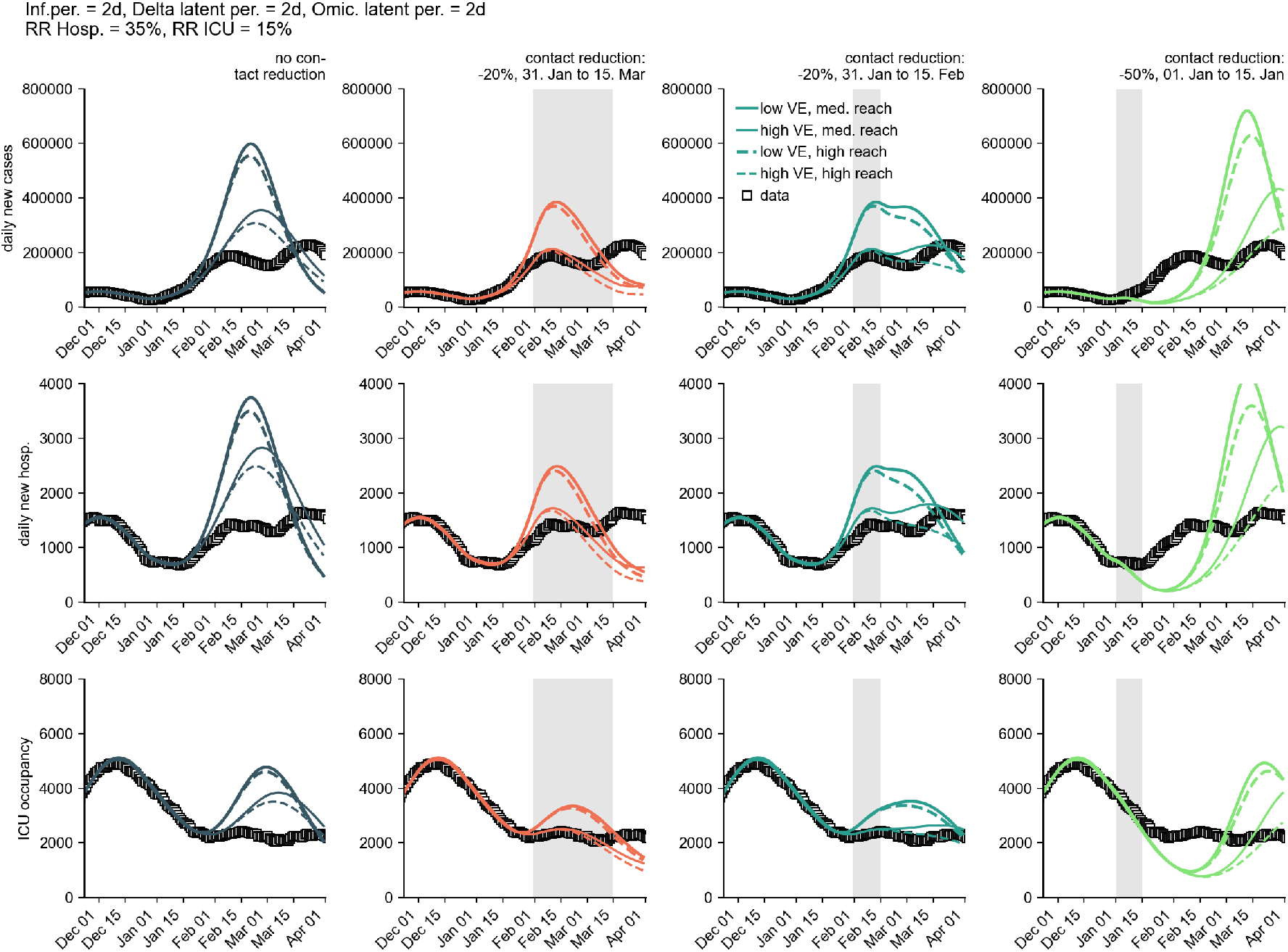
Comparison of model results for different contact reductions. An early, strong contact reduction could have lead to a strong rebound effect (right column). A slighter, long contact reduction (−20% by March 15) lead to a smaller outbreak (by April 1, second column from left). However, this could have been followed by another smaller wave after the end of the contact reduction period (not shown here due to uncertainties in the forecast horizon beyond March). A slight, short contact reduction led to a flattening of the infection wave and thus also to containment (second column from right) with a sustained continuation of systemic immunity by infection. Shown are results for the number of new cases per day, new hospitalizations per day and ICU occupancy for a combination of different model assumptions. Iterated here were “medium reach” and “high reach” of the booster campaign, “low VE” and “high VE” of the booster vaccination, as well as various contact reductions. Here, a generation time of 4 days was assumed for both variants (2 day latency). For each scenario combination, a simulation was performed with an average course of the contact behavior (stochastic simulation with zero variance). Furthermore, the relative risk of hospitalization by Omicron versus Delta was assumed to be *RR* = 0.35 and intensive care *RR* = 0.15.

**FIG. 5.**
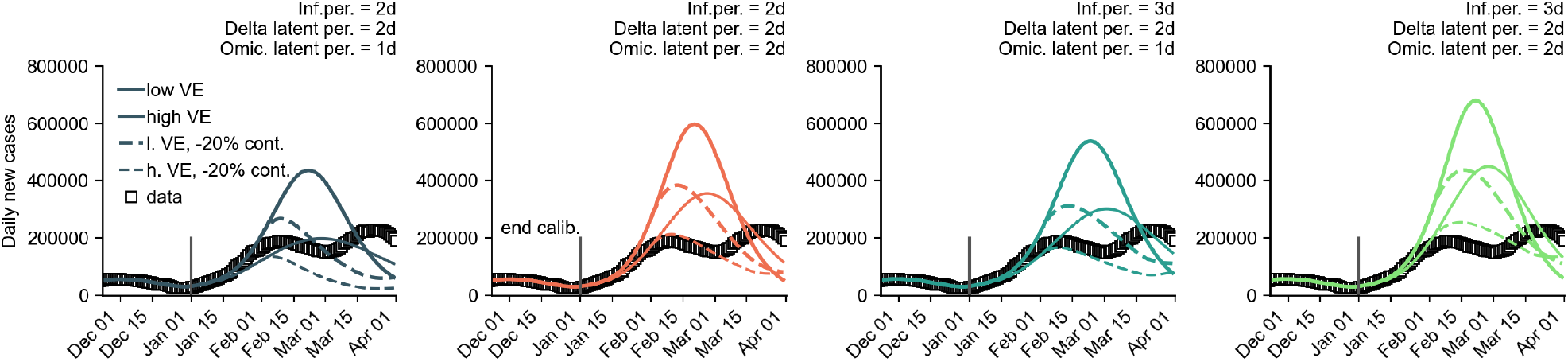
Contact reduction of −20% compared to the original trajectories for all generation times. Shown are results for the number of new cases per day for a combination of different model assumptions. “Medium reach” of the booster campaign, “low VE” and “high VE” of the booster vaccination were also iterated here. For each scenario combination, a simulation was performed with an average course of the contact behavior (stochastic simulation with zero variance). Furthermore, the relative risk of hospitalization by Omicron versus Delta was assumed to be *RR* = 0.35 and intensive care *RR* = 0.15.

For unchanged contact behavior, we found a maximum permissible relative risk (RR) of requiring intensive care in the range of 10%–20% in order to keep ICU occupancy below a critical value of 4 800 beds (see Tables III–XI).

Results for cumulative outbreak sizes and maxima of incidence, hospitalization incidence, and ICU occupancy are shown in Tables XI–XIV and Tables VII–X.

The hypothetical case of a short-term, drastic increase in the initial immunization rate illustrated the contribution to the pandemic of those who were still unvaccinated (see Fig. 6). Here, it was assumed that, as of January 22, 2022, 15 million previously unvaccinated individuals would have achieved the initial full immunization status. A high initial immunization rate would have resulted in a large reduction in ICU burden due to the high efficacy of the vaccines against severe courses.

**FIG. 6.**
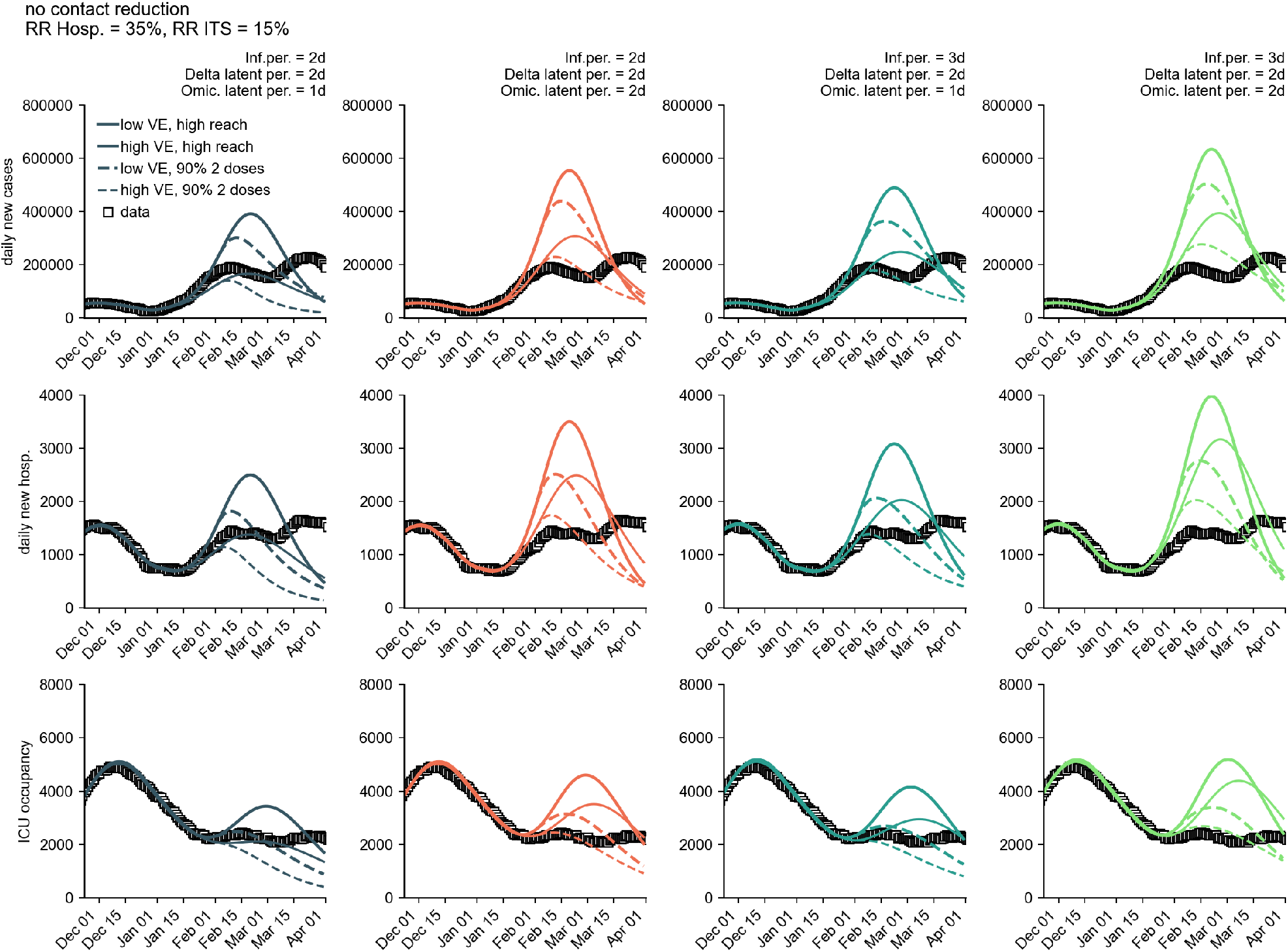
Comparison of model trajectories if, as of January 22, 2022, 15 million previously unvaccinated persons would have the same level of immune protection as after completion of the initial vaccination series. Shown are the number of new cases per day (first row), new hospitalizations per day (second row), and ICU occupancy (third row) for a combination of different model assumptions (see Methods). The solid curves correspond to the results of the “high reach” scenario from Fig. 1, the dashed curves to the corresponding scenario of an initial immunization rate of approx. 90%.

Retrospectively, our results agreed well with the scenario “medium reach”, “high VE”, and −20% contact reductions (short time in early February 2022), cf. third column from the left in Fig. 4. The “medium reach” assumption of administering booster vaccinations to 80% of people who had a completed first vaccination series proved to be an accurate estimation. Furthermore, the temporal evolution of the vaccine efficacy in the “high VE” scenario shows satisfying agreement with the time series of VE that was estimated using Farrington’s method counting breakthrough infections, see Fig. 15 (with the data containing mostly symptomatic infections, however, which hinders a definite and direct comparison). The short decrease in growth rate forced by a 20% reduction in the contact modulation led to a peak size of ≈ 200, 000 new cases per day in the model, followed by a resurgence and a slightly larger second peak, both of which reflects the actual time series of the outbreak quite well. Since no decrease in contact or mobility behavior was observed in Germany during this period of time, however, the first decrease of the growth rate has probably been induced by other factors and was likely caused by an incipient systemic immunity, which implies that we might have underestimated the initial systemic immunity against infection with the Omicron variant. The second peak was, with high certainty, caused entirely by the spread of the BA.2 sublineage of Omicron, which was associated with an even higher base transmissibility and therefore led to a net increase in growth rate after the initial drop. The combination of these two effects has likely led to a temporal evolution of the growth rate that was reflected well by the “−20% contact reduction for a short time” scenario in the model.

## 4. DISCUSSION

The presented results are subject to a number of limitations due to both model structure, as well as various uncertainties in the assumptions. For example, population-averaged models that do not explicitly distinguish between vaccinated and unvaccinated individuals can systematically overes-timate the size of major outbreaks by an order of magnitude of approximately 10%. We therefore expected the actual outbreak size to be lower than those reported here. Nevertheless, we decided to not explicitly make a distinction between vaccinated and unvaccinated individuals, enabling us to adjust, simulate, and reanalyze the model in a dynamical manner. This facilitated an agile response to changes in data—an advantage that justified uncertainties and allowed for sustainable analysis procedures due to the structural stability of the model. By comparing many different scenarios it was thus possible to quickly analyze and illustrate which aspects of the expected dynamics are robust to parameter changes and to which the model reacted sensitive to.

Further systematic overestimation of outbreak sizes may result from assumptions about the contact structure. In the present case, a homogeneous contact structure was assumed, which is the same within and between all age groups of the population. This simplification, which is not met in reality, likely leads to an overestimation of the number of cases since heterogeneities in the contact structure of age groups usually leads to lower outbreak sizes [19]. Furthermore, this effect may have led to an underestimation of the relative risk of hospitalization and requiring intensive care, as the dynamics at the beginning of a wave are often dominated by younger age groups, which are usually at lower risk of severe disease. As more elderly people become infected, for whom the probability of a severe course is higher than for younger people, the at-the-time observed relative risk may have increased again and, with it, ICU occupancy. However, as with the distinction in vaccination status, a heterogeneous contact structure was also disregarded in favor of reducing model complexity.

The average contact behavior in December 2021 was chosen as the basis for extending the contact modulation. At the time, it was reasonable to assume that this contact behavior could be extended into January 2022 and the following months due to then-unchanging protective measures. Since the contact behavior was not at pre-pandemic levels, it could be assumed that increased contact behavior would be observed by the end of the first Omicron wave at the latest, which could lead to another wave. Due to large uncertainties as to when an increased contact behavior could be expected, to what extent this increase would occur, uncertainties as to how long an Omicron infec-tion protects against re-infection, uncertainties regarding a possible underreporting of infections, as well as the influence of seasonality on the spread of Omicron, this effect was not considered here. In this sense, an underestimation of the number of cases beyond April 1, 2022 was to be expected.

Furthermore, the concept of a possible *emergency brake* was disregarded in this study, i.e., no automatic contact reduction were to be implemented as soon as case numbers, hospitalizations or ICU occupancy exceeded a critical value. However, we illustrated this effect by the influence of a long, weak contact reduction of -20%.

There are also other possible limitations due to uncertainties in the assumptions made or the processes underlying the model:

i. At the time of model development in mid-December 2021, there was uncertainty about the immunity of recovered individuals to reinfection with Omicron. In the model, all persons recovered from the first three pandemic waves were assumed to be susceptible (except vaccinated recovered individuals, who were equated with vaccinated persons that were not previously infected), while those who recovered from the Delta wave were assumed to be 100% immune. This results in an infection-induced initial systemic immunity that lies between these two extremes. In reality, increased immunity of those infected from earlier waves could lead to an overestimation of peak heights in the model. In the same sense, reduced immunity after a Delta infection or decreasing immunity over time, not taken into account here, could be the cause of an underestimation of the peak heights. Mathematically, our approach only served the purpose of assuming a certain, comparatively low, basic immunity against infection with Omicron in the population, i.e. the above-mentioned model assumptions cannot be easily transferred to reality. However, due to the unclear data regarding the immunity of recovered persons with respect to infections with Omicron during the development of the model in mid-December 2021, our approach can be considered feasible at the time.
ii. Similarly, there was uncertainty in the number of recovered persons who received a vaccination after infection and, thus, achieved at least primary immunization status. Since recovered individuals from the first waves were assumed to have no immunity (see above) and treated the same as susceptibles, we implicitly assumed a vaccination rate of recovered individuals equal to the population-wide vaccination rate. Given the unclear data situation during model development in mid-December 2021, this approach can also be considered practicable.
iii. With regard to the exact estimation of the expected number of cases, it should be noted that the assumed under-ascertainment (i.e., the proportion of unreported infections) was also subject to uncertainty and could only be roughly estimated. In the present case, we assumed a constant reporting rate of 50%, i.e., every second infection was reported. If the number of unreported infections was higher in reality, the observed case numbers were likely to be smaller than those estimated by the model because natural population-wide immunity would be achieved earlier at the then-current level of contact behavior (effective R-value of *R*_eff_ < 1). Similarly, underreporting may have further increased in the following months due to changes in prioritization of testing or other logistical limitations. Conversely, a relaxation of the situation could have led to a subsequent increase in ascertainment during the decline of an epidemic wave and therefore a decay in the reported number of infections that is slower than the true decline. At the time, however, a constant reporting rate of 50% seemed a plausible assumption [20].

The VE against *infection* with Delta assumed in this analysis was approximately equal to the VE against *symptomatic disease* with Delta, a result of regression of the collected data on VEs. However, because many studies estimated VE against infection to be lower than VE against symptomatic disease, we may have been overestimating the efficacy of vaccines against infection with Delta. Furthermore, ignoring the VE of the AstraZeneca and Johnson & Johnson vector vaccines also leads to a systematic, though likely not considerable, overestimation of the VE against infection with Delta. In the case of VE overestimation, Omicron growth would haven been driven more by an increase in base transmissibility, as immune evasion would be lower. Since higher transmissibility leads to greater outbreaks, this would have implied an underestimation of outbreak size in our results. However, the model also ignored VE against transmission, which has been observed to be non-negligible in several countries. This VE against transmission would again raise the effective contribution of vaccination to the attenuation of the incidence. In retrospect, we seem to not have overestimated the efficacy of vaccines to prevent infections with Omicron.

To sum up, we devised and analyzed a parsimonious infectious disease model that was able to capture the central aspects of the spread of the Omicron variant in Germany before it dominated the dynamics in early 2022, despite many uncertainties and limitations. We expect that our methodology can be used to evaluate future outbreaks, either caused by other emerging SARS-CoV-2 variants, or with regards to other infectious diseases.

## 5. FULL METHODOLOGY

### 5.1. Model definition

Infection dynamics follow a temporally-forced susceptible-exposed-infected-recovered (SEIR) model:

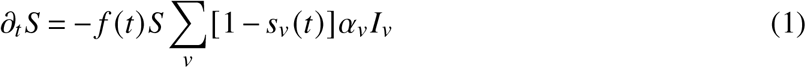

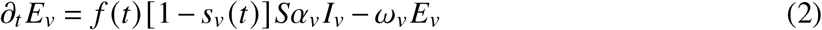

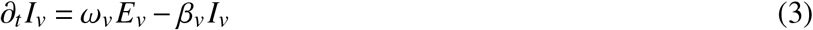

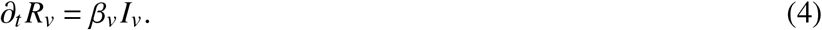

Here, *v* is a “variant of concern” (hereinafter “variant” or “VOC”), *f* (*t*) controls for time-varying contact behavior (e.g., through non-pharmaceutical interventions (NPIs) or voluntary behavioral change), *s*_*v*_ (*t*) is the population-averaged efficacy of vaccination against infection (averaged across unvaccinated and vaccinated subpopulations). The transmissibility *α*, mean latency 1/*ω* and mean infectious period 1/*β* are potentially variant dependent.

To fit the model to the data, we add compartments for (i) reported, (ii) hospitalized, or (iii) in need of intensive care. Let *p*_*C*,*v*_ denote the probability of appearing in the reporting statistics after infection (compartments *C*), *p*_*H*,*v*_ the probability of an unvaccinated person being hospitalized after infection (i.e., becoming “severely” ill with COVID-19, compartments *H*) and *p*_*U*,*v*_ the likelihood of an unvaccinated individual requiring intensive care following infection (compartments *U*). Furthermore, *h*_*v*_ (*t*) and *u*_*v*_ (*t*) are time-dependent functions that quantify population-wide vaccine efficacies against hospitalization and intensive care.

To adequately reflect the respective empirical distributions, we assign Erlang distributions with number *n*_•_ and rate *n*_•_/*τ*_•_, (i.e., distributions with mean *τ*_•_ and standard deviation 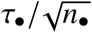) to the transition times between infection and reporting, hospitalization, and ICU admission.

Thus, we obtain

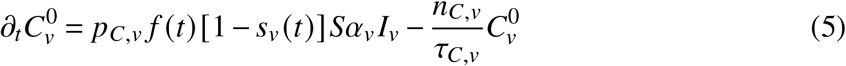

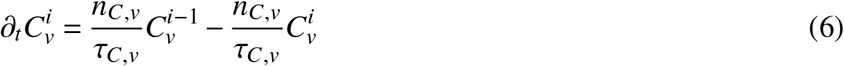

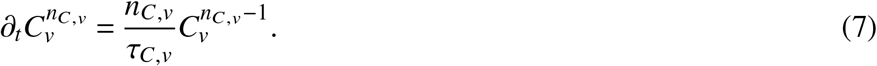

The variant-dependent incidence is given by

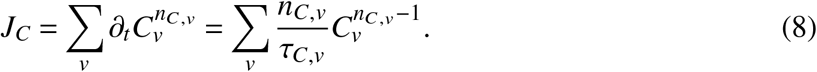

For hospitalizations, we define

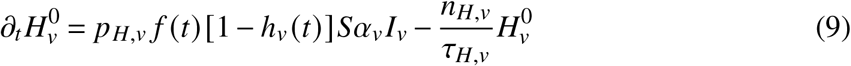

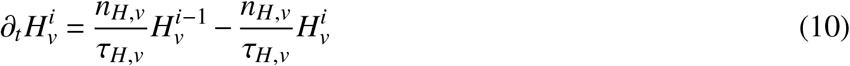

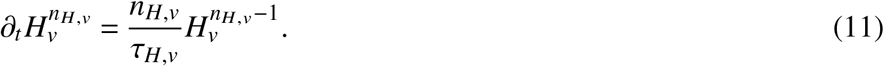

Here, *h*_*v*_ (*t*) is the population-averaged efficacy against hospitalization at time *t*. The number of new hospitalizations at time *t* is

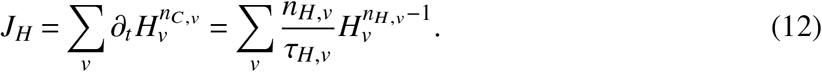

To model the number of ICU beds occupied, we define distributions for both the transition between infection and ICU admission and the length of stay in ICU. The equations follow

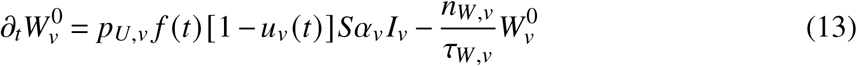

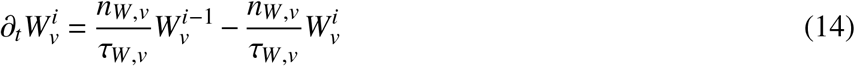

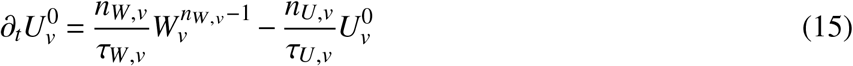

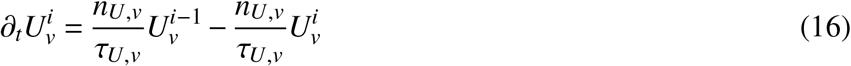

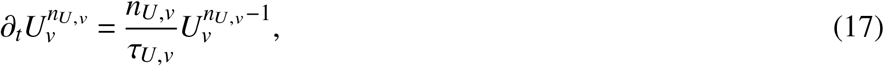

with a total intensive care unit occupancy

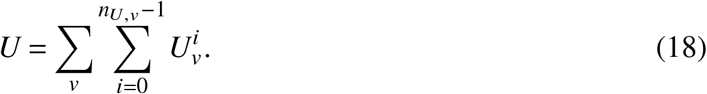

### 5.2. Population-wide vaccine efficacies

To find time-dependent population-wide vaccine efficacies (“VEs”) against infection *s*_*v*_ (*t*), hospitalization *h*_*v*_ (*t*) and requiring intensive care *u*_*v*_ (*t*), we assume four basic vaccination states: VE 14 days after completion of first vaccination series (subscript “2”), VE after waning, subscript “2, *w*”, VE 7 days after booster vaccination (“booster”, subscript “*B*”), and booster VE after waning (subscript “*B, w*”. We define the mean time of VE attenuation as *θ*_2_ and *θ*_*B*_. Initially, the entire population is unvaccinated, *A* = 1. We model the temporal transition from unvaccinated to vaccinated of different statuses *V*_•_ for each variant as

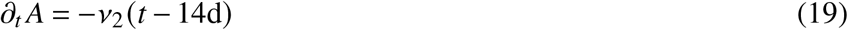

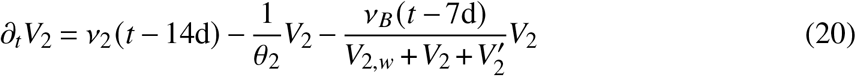

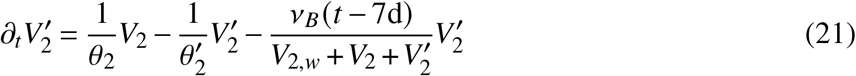

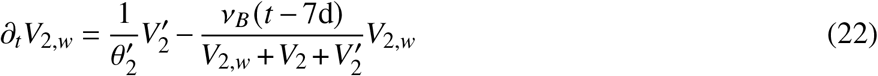

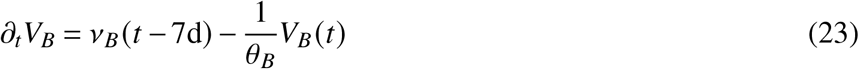

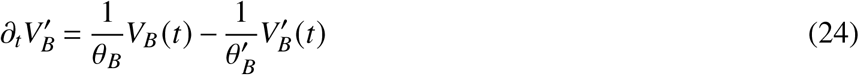

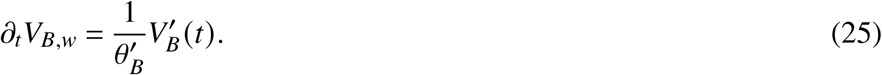

The intermediate compartments 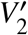 and 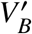 ensure a realistic approximation of the actual waning time distributions. Here, we use the rates of completed vaccinations *v*_2_(*t*) and booster vaccinations *v*_*B*_ (*t*), which can be determined as the progressive differences of the respective cumulative number of vaccinations [21]. Note that we assume the onset of maximum protection at 14 days after completion of the first vaccinations series and at 7 days after booster vaccination. *V* and *V*^′^ represent states of maximum immunity that were constant for a mean time 2*θ* and decreased thereafter (state *V*_*w*_) at rate 1/*θ*^′^.

Let *e*_*v*,•_ be a placeholder for different VEs against infection with variant *v* and • be a place-holder for vaccination statuses “2 doses” and “boostered”, as well as initial and “waned”. The population-averaged VE with respect to variant *v* is calculated as

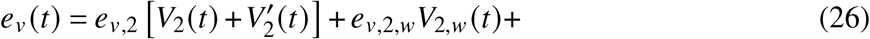

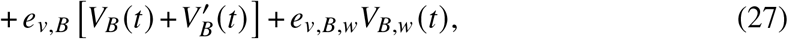

and the VE for vaccinated-only individuals

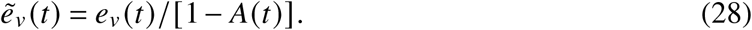

Here, *e*_2,*v*_ is the maximum VE 14 days after completion of the vaccination series and *e*_2,*w*,*v*_ is the minimum VE reached after a mean time of 2*θ* _•_.

With 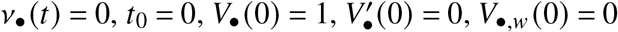, an average sigmoidal decrease in VE from the time of maximum immunity is obtained from the above equations to be

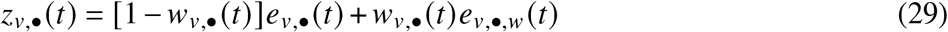

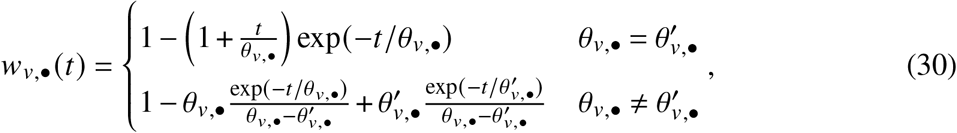

using the values of *e*_•_ and *θ*_•_ derived from data fits.

To estimate the temporal decay of different VEs after two doses against infection, against symptomatic COVID-19, or against a severe course with variant Delta, we used data from the meta-review by Feikin *et al*. [22], only considering data explicitly reporting VE against Delta. Time intervals since the administration of the second vaccine dose and the computed VE within these intervals were extracted. The mean of each interval minus 14 days was used to define the day of VE since peak immunity after two doses. If only the beginning of a time interval was defined (e.g., starting 140 days after administration), the length of the time interval was measured from the previous one (e.g., time interval 1: “days 105 to 139”, time interval 2: “days 140+”; time interval 2 was then assigned an upper bound of (140d + 139d − 105d) = 174d). One study (Tartof *et al*.[23]) identified in the review by Harder *et al*. [24] refers to an additional time point that was also considered here. Also mentioned in Harder *et al*. is a study by Fowlkes *et al*.[25], from which data of VE against infection with Delta after 2 doses were extracted. Here, too, incomplete time intervals were fitted in a manner analogous to Feikin *et al*.. Andrews *et al*.[26] provided data on VE against symptomatic infection with both Delta and Omicron after 2 doses as well as after booster vaccination. Because this study specifies time intervals in weeks, it was assumed that the first interval (week 2-9) corresponded to days 14 to 63 and subsequent intervals (weeks 10-14, 15-19) each corresponded to the next day of the previous interval until the end of the last week in the interval (e.g., week 10-14 thus corresponded to day 64 to 98). The same procedure was used for booster vaccination data and was implemented here as well. Unless otherwise defined in the study, the mean of each time interval minus 7 days (rather than 14 days) was used to define the day of VE since maximum immunity. Data on VEs against hospitalization and intensive care with Delta were taken from a study by de Gier *et al*. [27]. The UK Health Security Agency published data on VE against symptomatic infection with Delta and Omicron and against hospitalization with Omicron after two vaccine doses and after one booster [7]. Data on VE against infection with Delta and Omicron were extracted from Hansen *et al*. [13]. Because protection after booster vaccination was determined here only as a comparison with fully vaccinated persons, only data on VE after 2 doses were extracted. In addition, the 95% confidence intervals of VEs were taken from all studies and used as weights in fits. Our study exclusively refers to data published by the end of 2021.

Eq. (29) was fitted to these data per variant, vaccination status, and target variable to obtain values for *e*_*v*,•_, *e*_*v*,•,*w*_, *θ*_*v*,•_ and 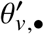. Assumptions were made for VEs for which no data are available. The time courses of average VEs obtained by this procedure are shown in Fig. 2.

For the future course of the vaccination campaign, we assumed that no more initial vaccinations were administered. Furthermore, we assumed that the rate of daily booster vaccinations maintained its level achieved in late December, such that the cumulative number of booster vaccinations, following a sigmoid function, reached (i) the number of all initially vaccinated persons in 2021 and (ii) 80% of those (see Fig. 7).

**FIG. 7.**
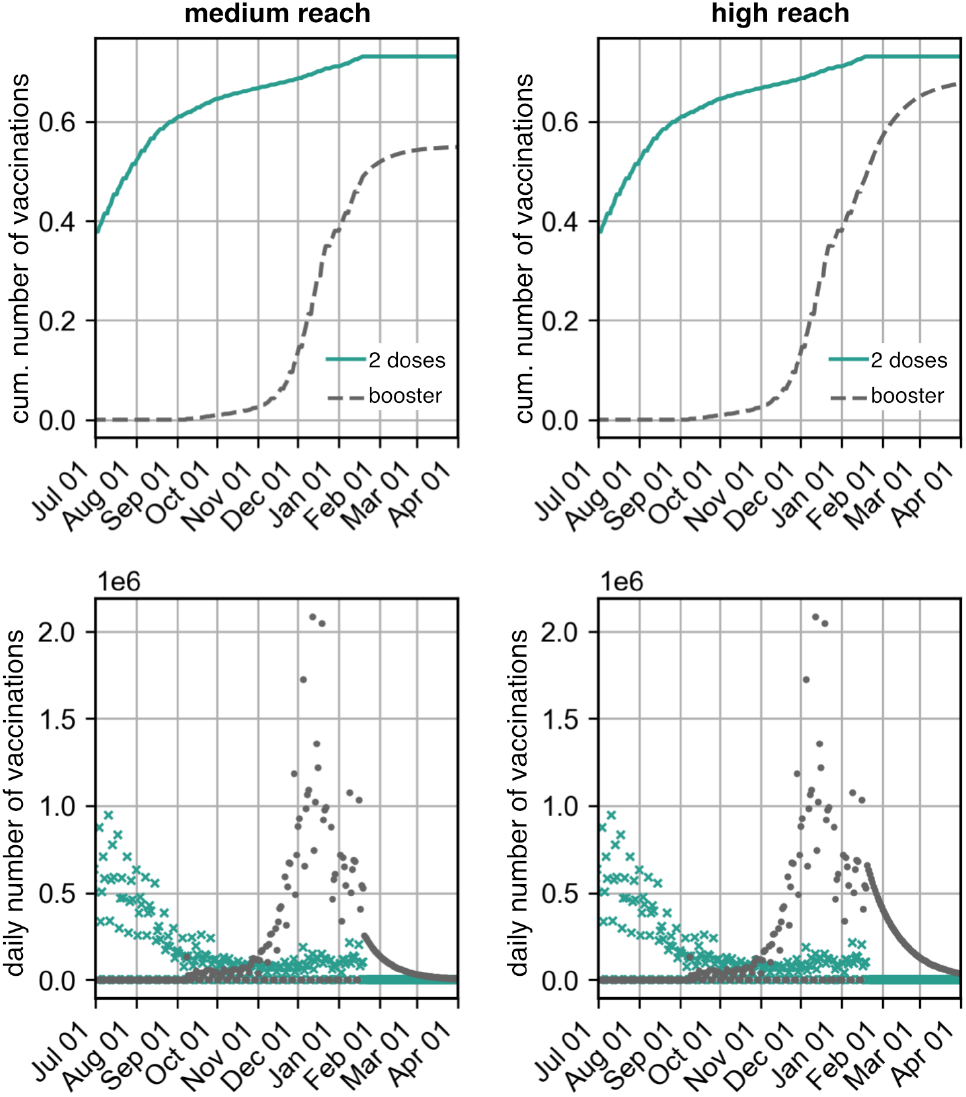
Cumulative number of vaccinated persons and number of daily vaccinations (data and extrapolation) assuming that the number of booster vaccinations reached 80% (medium reach) or 100% (high reach) of those first vaccinated by the end of 2021.

With these fits and assumptions, Eq. (19) was integrated to obtain the population-averaged VEs in Eq. (26), shown in Figs. 8–9.

**FIG. 8.**
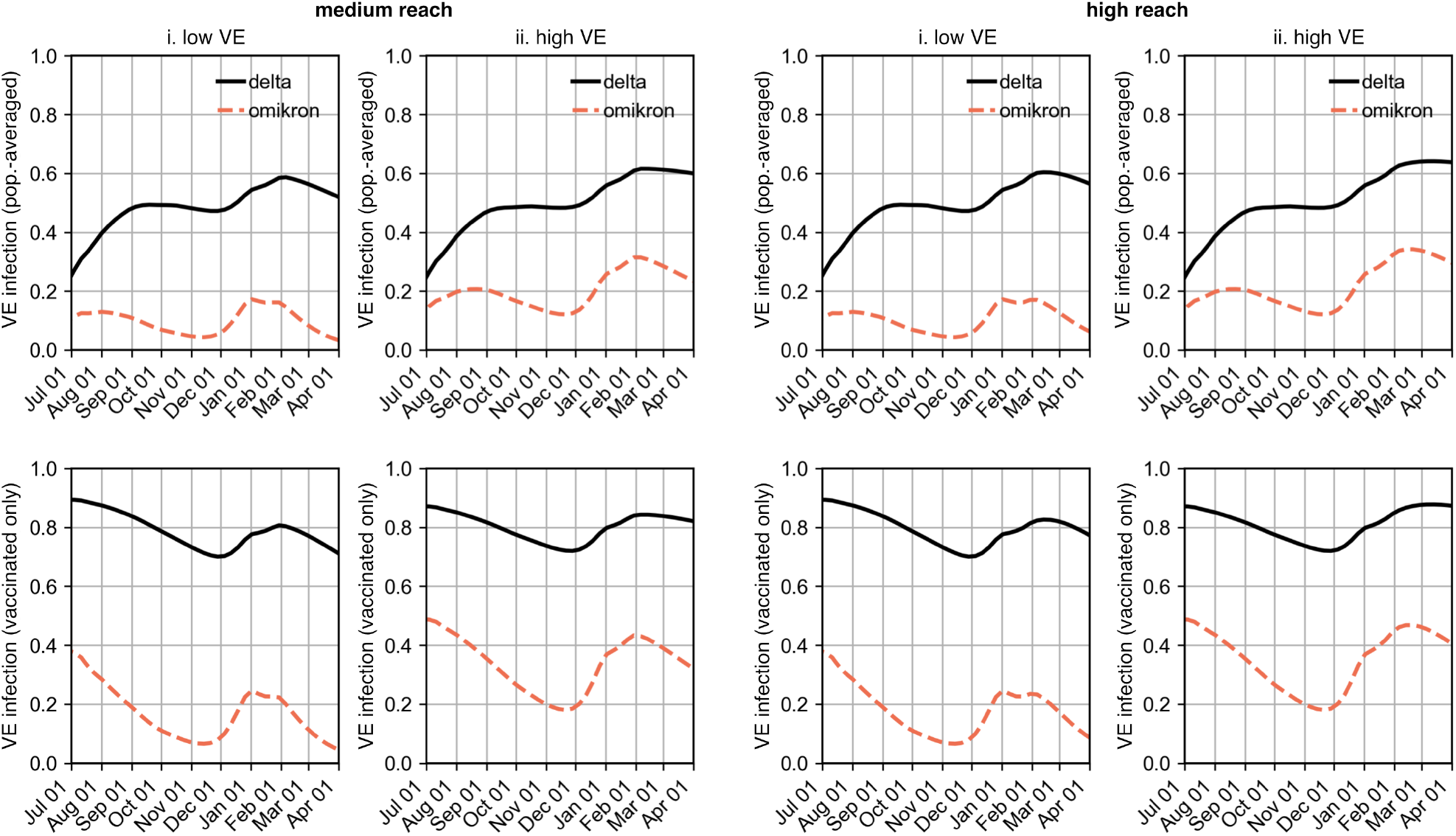
Population-wide VE (top row) and VE of the vaccinated population (bottom row) against infection under the assumed vaccination efficacies from Fig. 2. (i) Results according to the data for VE against infection of first-immunized (low VE) and (ii.) assuming that VE against infection matched the data for VE against symptomatic infection (high VE). Both scenarios (low VE and high VE) were each considered under the assumption that either 80% (medium reach) or 100% (high reach) of those first immunized by the end of 2021 received booster vaccination.

**FIG. 9.**
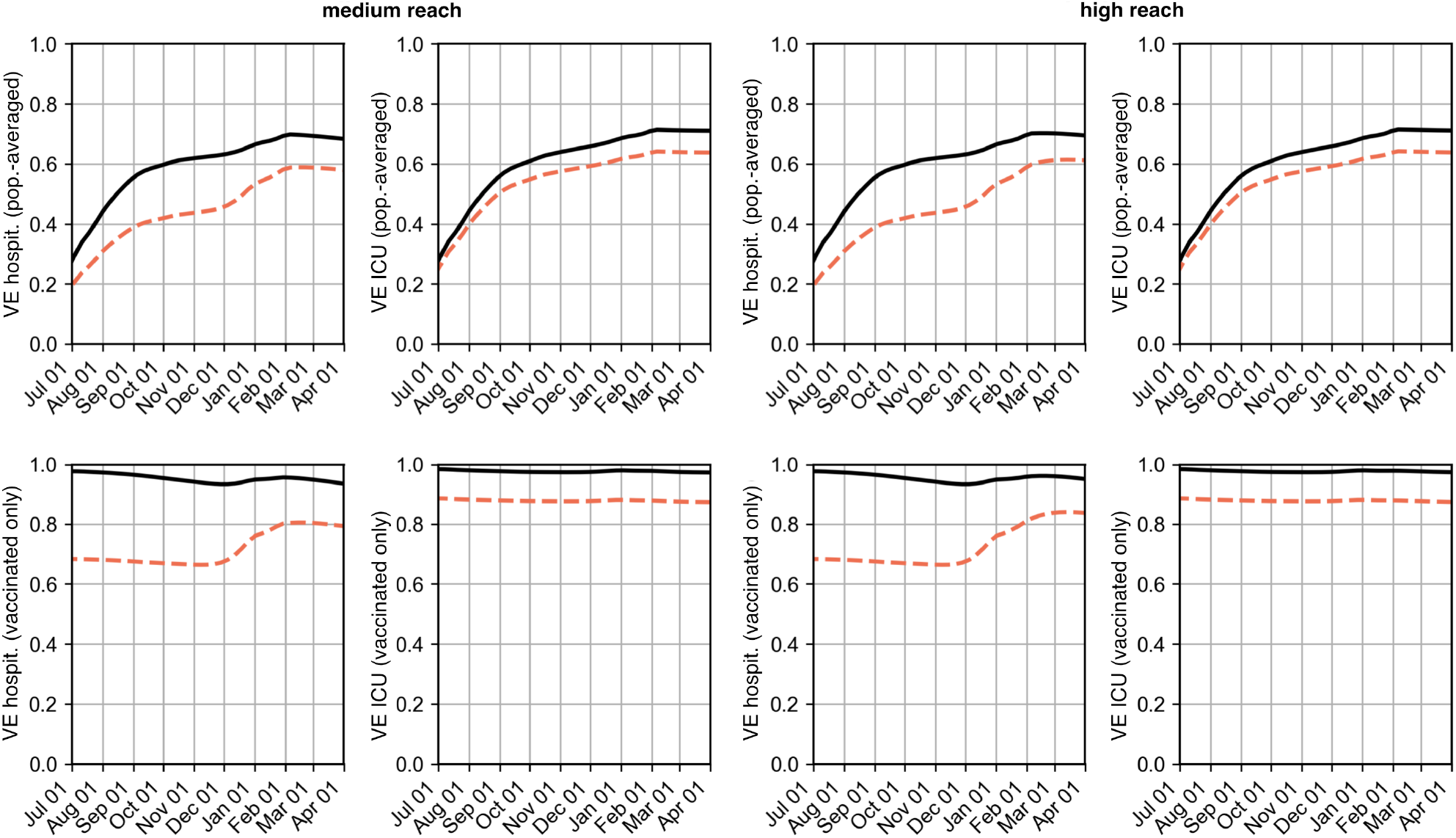
Population-wide VE (top row) and VE of the vaccinated population (bottom row) against hospitalization and intensive care under the assumed vaccine efficacies from Fig. 2. Both scenarios (low VE and high VE) were each considered under the assumption that either 80% (medium reach) or 100% (high reach) of those first immunized by the end of 2021 receive booster vaccination.

### 5.3. Calibration of transmissibility of VOCs to growth rates in December 2021

To determine the transmissibility of the VOCs Delta and Omicron in Germany, we measured the respective growth rates of the variants in December 2021 using reported data and data from the German Electronic Sequence Data Hub (DESH) and applied analytical approximations to derive the transmissibilities.

The growth rate Λ_*v*_ of a variant *v* at time *t* is given by the largest eigenvalue of the Jacobi matrix of the ODE system from Eqs. (1–4) as

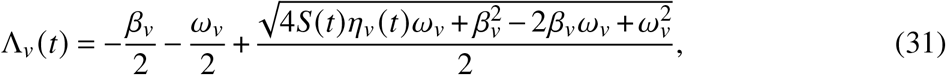

where *η*_*v*_ (*t*) = *f* (*t*)*α*_*v*_ [1 − *s*_*v*_ (*t*)] is the time-dependent infection rate of a variant modulated with time-dependent contact behavior *f* (*t*). Here, *S*(*t*) is the time-varying relative proportion of susceptibles and *s*_*v*_ (*t*) was the population-wide vaccine efficacy against infection. From Eq. (31), the base transmissibility of a variant is given by

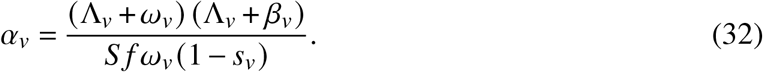

From the fixation dynamics of Omicron (Fig. 10), a fit of the function 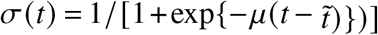 to the measured proportion of Omicron in all new infections can be used to determine the fixation rate *μ*, which is related to the growth rates of the variants as

**FIG. 10.**
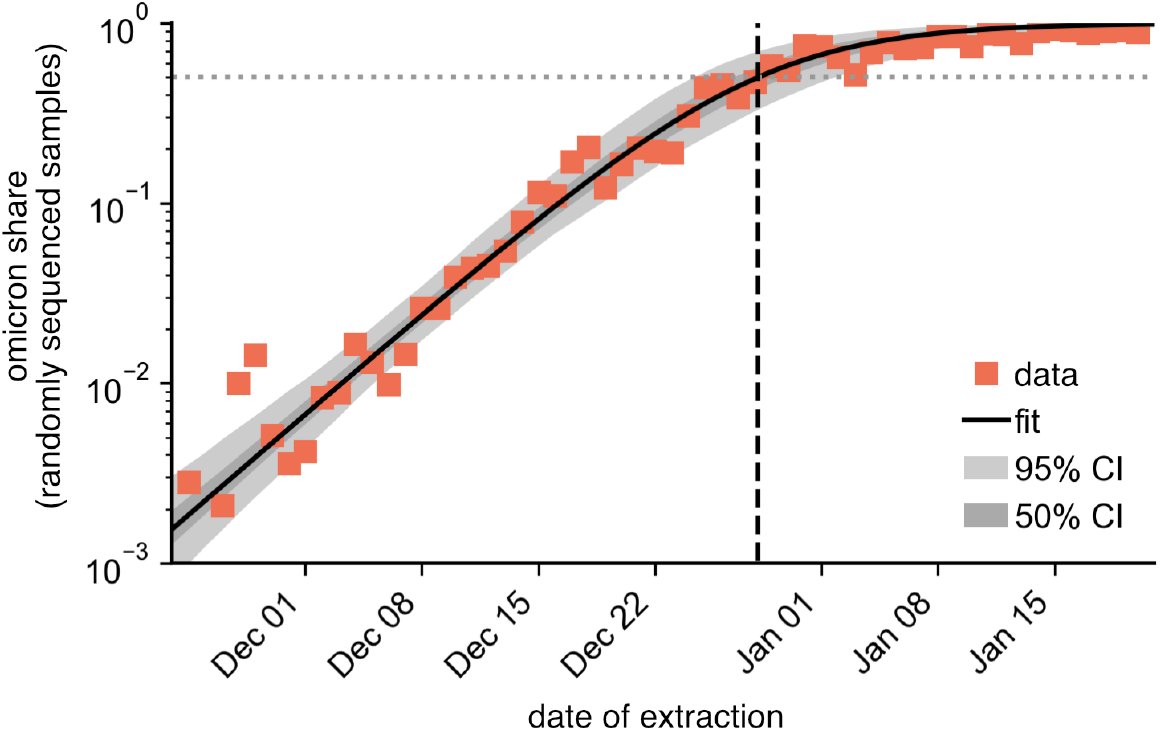
Omicron fixation dynamics with numerical fit (sigmoid function, see main text). Data points represent the Omicron proportion in the random laboratory sample by date of extraction [33]. The dashed vertical line marks the time at which the Omicron proportion was 50% according to the fit.

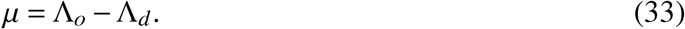

Hence follows

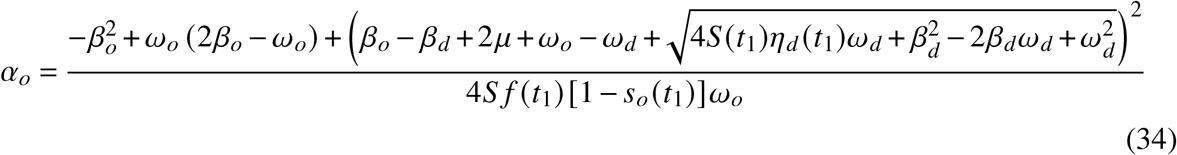

on a calibration date *t*_1_.

The proportion of new infections attributed to the Omicron variant was measured from random laboratory samples of DESH data [28]. We assumed that the sample collection date corresponded to the date of symptom onset. We filtered collected samples by the “scorpio_call” column, in which a sequenced genome is assigned to a VOC using the “Scorpio” software (see descriptions in [29–31]). Here, all sequences whose “scorpio_call” value contained the string ‘Delta’ were assigned to the VOC Delta and all sequences whose “scorpio_call” value contained the string ‘Omicron’ were assigned to the VOC Omicron (this included the value ‘probable Omicron’). In addition, filtering was done for sequences that were randomly selected for sequencing resulting in ℵ_*d*_ (*t*) sequences of VOC Delta and ℵ_*o*_ (*t*) sequences of VOC Omicron for each day. The proportion 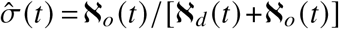 could then be fitted to the function 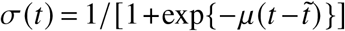 with free parameters *μ* and 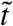 (time at which Omicron would account for 50% of new infections). The fit was performed with MCMC sampling to minimize the sum of residuals in logarithmic space 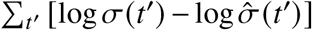, with 100 walkers and 1000 steps each. We thus obtained an ensemble of 100 000 parameter pairs 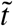 and *μ*. We found mean values of ⟨*μ*⟩ = (0.184 ± 0.019)d^−1^ and 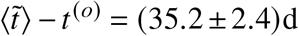, where *t*^(*o*)^ = November 23, 2021 (date of collection of the first Omicron samples).

To determine the growth rate of Delta, we used the number of new infections after symptom onset, imputed using a nowcasting technique [32] and restricted ourselves to the period between December 1, 2021 and December 15, 2021. Let *Ĵ*_*S*,tot_ (*t*) be the number of new infections after symptom onset on date *t*. Then *Ĵ*_*S*,*d*_ (*t*) = *Ĵ*_*S*,tot_ (*t*)*σ*(*t*) is the number of daily new infections with Delta (with mean values of *μ* and 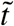). In this way, we transformed the measured total new infections and fit an exponential decrease *J*_*s*_ (*t*) to *Ĵ*_*S*,*d*_ (*t*).

The doubling time of a variant was measured in the model as

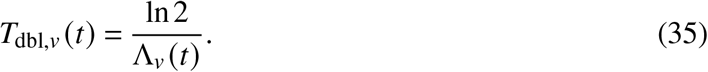

In the analysis, this resulted in 4.5 to 5.5 days for Omicron cases in December 2021.

By Eq. (32), the ratio of the base transmissibility of two variants is given as

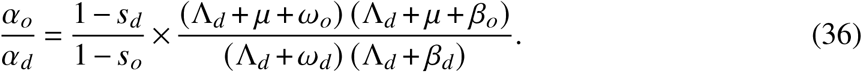

In the special case of constant latency and infectious period, as well as *s*_*d*_ *≡ s* and *s*_*o*_ *≡* (1 − *ϵ*)*s* with “immune evasion” *ϵ*, we find

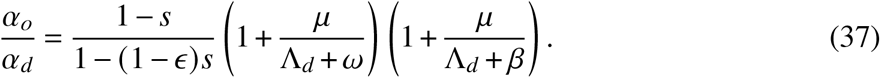

From this equation, it can be seen that both smaller latency periods (larger *ω*) and smaller infectious periods (larger *β*) require smaller increases in base transmissibility to explain the observed rates Λ_*d*_ and *μ*. For example, with *s* = 0.5, *ϵ* = 0.9, *μ* = 0.19/d, Λ_*d*_ = −0.045/d, and *ω* = 1/2d, resulting in an increase of *β*_1_ = 1/7d for (*α*_*o*_/*α*_*d*_)_1_ = 100% + 119% and an increase of *β*_2_ = 1/3d for (*α*_*o*_/*α*_*d*_)_2_ = 100% + 24%.

### 5.4. Calibration of the contact modulation

The transmissibility of Delta *α*_*d*_ was chosen such that ℛ_0_ = *α*_*d*_/*β*_*d*_ with fixed ℛ_0_. The exact value of ℛ_0_ played a minor role due to the freely chosen contact modulation *f* (*t*) (only the scale of *f* (*t*) was determined by ℛ_0_—a high ℛ_0_ required a lower *f* (*t*) to explain the observed rates than a lower ℛ_0_). The contact modulation at time *t* was derived from Eq. (31) to be

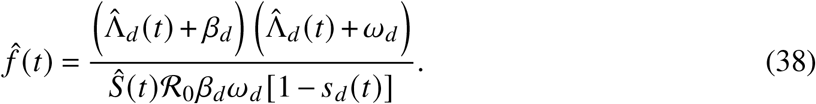

We determined the empirical growth rate by the growth of the incidence of Delta cases *Ĵ*_*C*,*d*_ = [1 − *σ*(*t*)] *Ĵ*_*C*_ (here, *Ĵ*_*C*_ is the 7-day average of total incidence by reporting date and *σ*(*t*) is the fitted fixation curve of the VOC Omicron), obtained from the identity Λ_*d*_ (*t*) = *∂*_*t*_ln*Ĵ*_*C*,*d*_ by

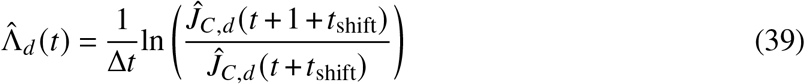

in the discrete approximation with Δ*t* = 1d and with reporting delay *t*_shift_. A value of *t*_shift_ = 7d ≈ *n*_*C*_*τ*_*C*_/(*n*_*C*_ + 1) satisfactorily approximated the reporting delay in practice. For the number of susceptibles at time *t*, we chose

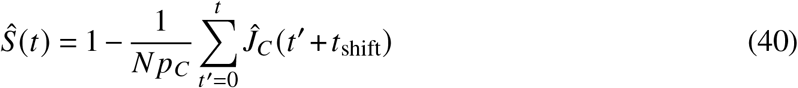

with the proportion of recorded cases *p*_*C*_.

The remaining free parameter for model calibration was the proportion of initially infected *I*_0_. We used the values shown in Tab. II.

### 5.5. Extrapolation of the contact modulation

Similar to the procedure in [**?**], we assumed that the empirically found contact modulation 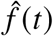 according to Eq. (38) followed a stochastic process with autocorrelation time *ϑ*^−1^ and extrapolated the series based on an Ornstein-Uhlenbeck process as

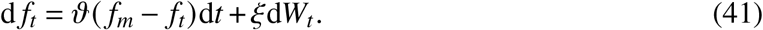

The process generates a time series *f*_*t*_ with mean *f*_*m*_ and variance *ξ*^2^/(2*ϑ*), using a Wiener process d*W*_*t*_. The autocorrelation time was obtained from the empirical curve 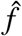 as approximately *ϑ*^−1^ = 21d. We chose *t*_end_ = 184d (January 1, 2022) as the start of the extrapolation time. To determine *f*_*m*_ and *ξ*, we measured the mean 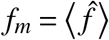 and the variance Var[*f*] in the period *t ∈* [150d, 184d]. The initial condition at time *t*_end_ was set to 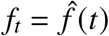, then the equation was integrated with Δ*t* = 0.1d and sampled with Δ*t* = 1d. The choice of the calibration end date had an impact on the course of the wave in early January 2022. Since the model course with *t*_end_ = 184d satisfactorily reflected the empirical data in January 2022, this value was not changed retrospectively.

The continuous function *f* (*t*) required for model integration was obtained from linear interpolation of the tabulated values of 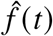 and *f*_*t*_ (tabulated for individual days).

Example runs for individual simulations are shown in Fig. 11.

**FIG. 11.**
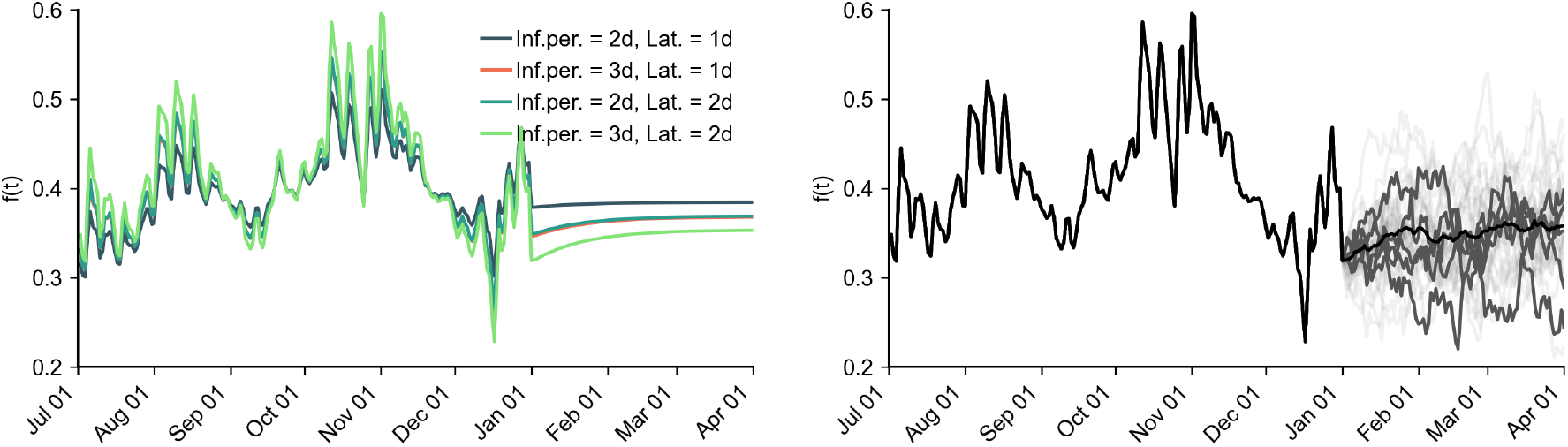
Time courses of contact modulation. (Left) Contact modulation for different generation times, inferred until Jan 1, 2022, then deterministically continued according to Eq. (41) with *ξ* = 0 (all curves for “medium reach” and “low VE”). (Right) Exemplary stochastic extrapolations of contact modulation according to Eq. (41) for “medium reach”, “low VE” and a generation time of 5 days (2 days + 3 days). The black curve shows the mean, dark gray curves show five randomly selected trajectories, and light gray curves show additional trajectories.

### 5.6. Latency and infectious period

The time scales relevant for SEIR models are latency *T*_*L*_ = 1/*ω* and infectious period *T*_*I*_ = 1/*β*, which sum to generation time *T*_*G*_ = *T*_*L*_ +*T*_*I*_ [34, 35].

We chose the latency period of VOC Delta as *T*_*L*_ = 2d, resulting from an incubation period of approximately 4 days [36] and the observation that the infectious period for the wild-type began, on average, 2 days before symptom onset [37]. One study from the UK [38] assumed a latency period of 2.5 days for both the wild-type and the VOC Alpha.

Another study from the UK found a mean generation time of approximately 5 days for VOC Delta [39], composed of a latency period of approximately 1 day, a presymptomatic infectious period of approximately 3 days, and a symptomatic infectious period of 1 day. To correspond to the generation time found in this way without changing the assumptions regarding latency period, we assumed an infectious period of 3 days (2 days pre-symptomatic and 1 day symptomatic) as a lower limit. In previous analyses for Germany, a shorter generation time of 4 days was assumed for VOC Delta, among others, which we have therefore also included in our analyses as a plausible value [32].

Due to limited data, we assumed in a first analysis that the VOC Omicron was associated with the same values for *T*_*L*_ and *T*_*I*_ as the VOC Delta. However, initial observations suggested that Omicron has a shorter serial interval (2.2 days on average in the Republic of Korea [40]) than Delta (3 days on average in the Republic of Singapore [41]). We therefore assumed for additional analyses that Omicron has a shorter latency of only *T*_*L*_ = 1d.

### 5.7. Remaining parameters

The number and immunity of recovered individuals was, at the time of analysis, unclear. A substantial number of recovered individuals was expected to have been vaccinated, and ergo part of the vaccinated population. We assumed that the immunity of the recovered decreases over time. Therefore, we calibrated the model to follow the Delta wave in the fall of 2021 and assumed that those recently recovered had full immunity against infection with Omicron, but that the unvaccinated recovered population from the first three pandemic waves had no protection against infection with Omicron, unless they had been vaccinated additionally. For this reason, we implemented the model starting July 1, 2021, with no recovered individuals initially.

We assumed a reporting rate of 50%, i.e., that every second infection was reported. To fit the model to the 7-day average incidence, we chose an Erlang distribution with *n* = 3 and *τ*_*C*_ = 11d between infection and reporting. Here, we mapped the incubation period of approximately 4 days, plus a reporting delay of 4 days [32] in addition to a 3-day systematic shift by the moving average. We took the 7-day average of new infections per day from ref. [33].

For the number of daily new hospitalizations, we assumed a hospitalization probability of *p*_*H*,*d*_ = 2.0% for unvaccinated persons, and a probability of intensive care of *p*_*U*,*d*_ = 0.45% (both values were per reported case, not per infection). We set the length of stay in an ICU equal to the observed length of stay observed during the first pandemic wave at *τ*_*U*_ = 18d and set *n*_*U*_ = 3 to represent the observed median and IQR sufficiently accurately [42]. Length of stay correlated strongly with ICU probability, so different pairs of values of these two parameters could generate rather similar trajectories of ICU occupancy. We chose the transition time from infection to hospitalization as *τ*_*H*_ = 13d with *n*_*H*_ = 2 and the transition time between infection and intensive care as *τ*_*W*_ = 14d with *n*_*U*_ = 1. We took the 7-day average of new hospitalizations per day from ref. [43] (adjusted time series) and the ICU occupancy in Germany from ref. [44]. The above values were chosen in a way that the model curves depicted the course of the Delta wave well (see as an example Fig. 12).

**FIG. 12.**
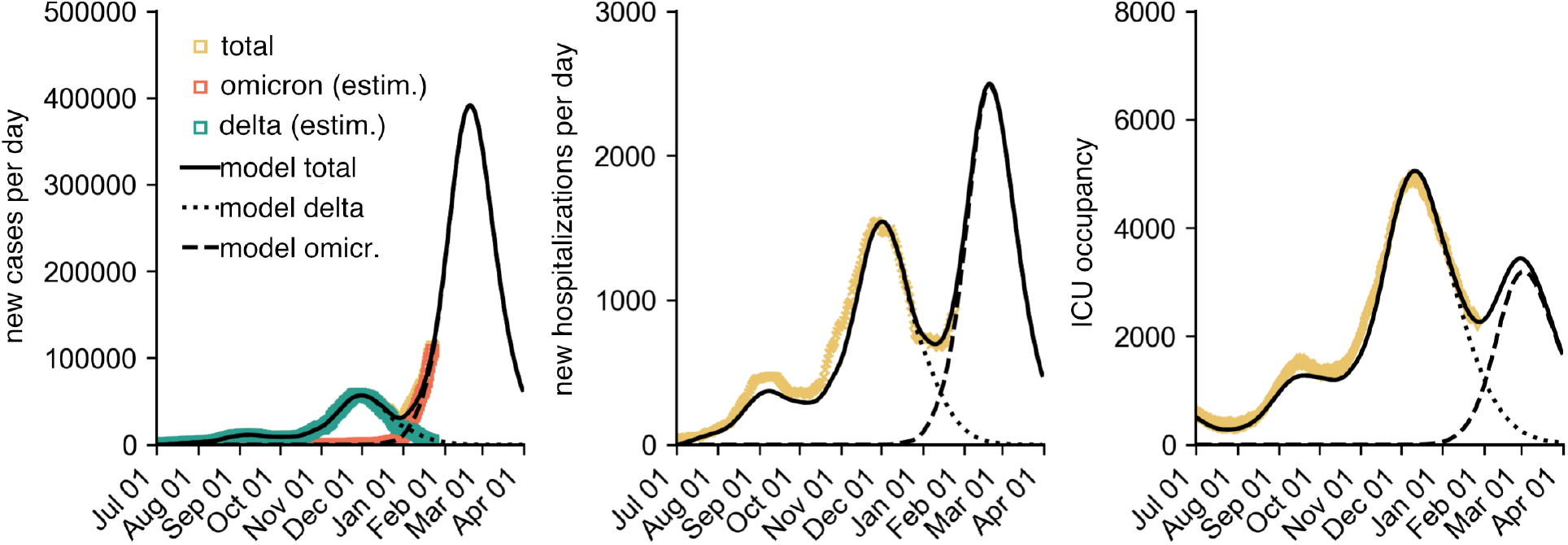
Calibration of the model to the fall of 2021 Delta wave assuming a mean latency and infectious period of 2 days each for Delta and 1 day and 2 days, respectively, for Omicron. This calibration further assumed that all of those fully vaccinated by the end of 2021 received a booster vaccination (high reach) and that VEs match the data in Fig. 2. In addition, the relative risk of hospitalization due to Omicron versus Delta was assumed to be *RR* = 0.35 and intensive care *RR* = 0.15.

We set *N* = 83155031 as the total population [43].

### 5.8. Contact reduction

To investigate the influence of different contact reductions on the course of the wave, we simulated 7 different scenarios. In the base scenario, contact modulations *f* (*t*) were not changed. For reductions, the curve was scaled between two time points *t*_*r*,0_ and *t*_*r*,1_. This resulted in the modified contact modulation

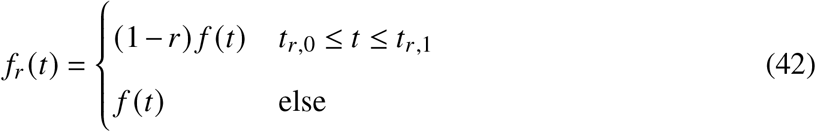

with 0 *≤ r ≤* 1. We chose values of (i) *r* = 20%, from January 31 to March 15, (ii) *r* = 50%, from January 31 to February 15, (iii) *r* = 50%, from January 31 to February 28, (iv) *r* = 50%, from January 31 to March 15, and (v) *r* = 50%, from February 15 to March 15. Example model runs for these contact reductions are shown in Fig. 13.

**FIG. 13.**
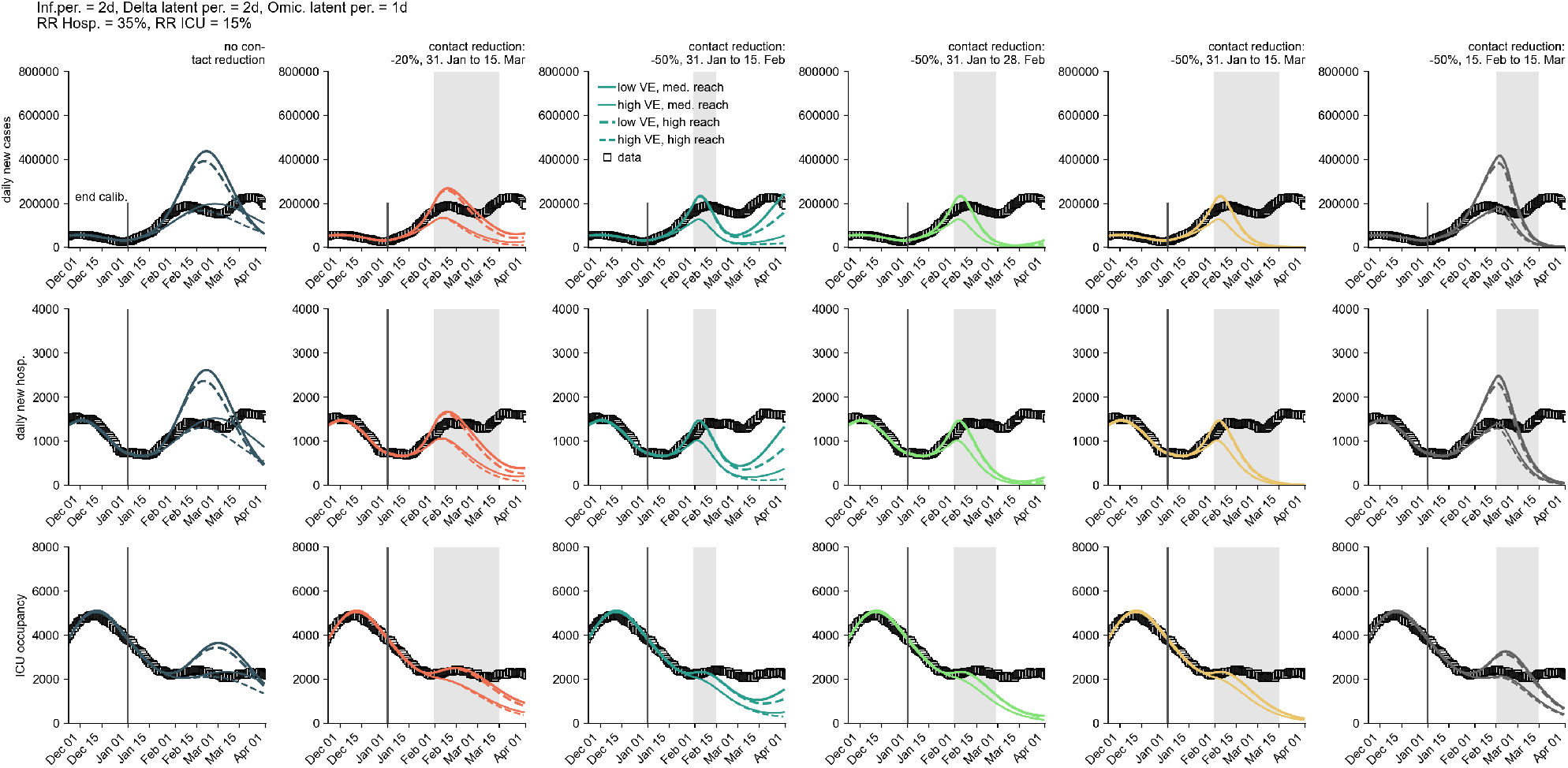
Different model runs for further contact reduction assumptions with a generation time of 4 days (2 days + 2 days). Intervals shaded in gray indicate the modeled periods of contact reductions.

### 5.9. Model simulations

The model was implemented and analyzed using the simulation software *epipack* [45]. As initial conditions for *t*_0_ = July 1, 2021, we chose the values shown in Table II. The model was integrated up to *t*_1_ = December 1, 2021, using a Runge-Kutta 4(5) method with dynamic step size control. We assumed an initial Omicron share of *σ*(*t*_1_) and fixed the modified initial conditions to 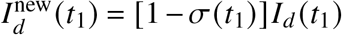 and 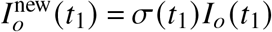 (all other compartments were assigned the respective values they assumed in the final state of the previous integration). Finally, the model was integrated until *t*_2_ = April 1, 2022. An example integration including the calibration based on the Delta wave is shown in Fig. 12).

**TABLE II.** Initial model conditions for *t*_0_ = July 1, 2021 for different values of latency and infectious period.

| Latency $\omega^{-1}$ [d] | Inf.per. $\beta^{-1}$ [d] | $I_0/10^{-5}$ |
| --- | --- | --- |
| 1 | 2 | 5.8 |
| 2 | 2 | 7.2 |
| 1 | 3 | 7.7 |
| 2 | 3 | 9.0 |

### 5.10. Overestimation of outbreak size

The model we devised for our main analysis does not explicitly distinguish between vaccinated and unvaccinated individuals, which reduces methodical complexity and facilitates quick adaption to new data as well as the derivation of analytical results. Yet, doing so can lead to a systematic overestimation of the outbreak size up to 10%. We illustrate this overestimation by comparing two toy models that neglect waning immunity, calibration to data, or contact modulation (see Fig. 14). Note that in SIR-like models, final outbreak size is independent of any latent compartments, which we therefore omit entirely.

**FIG. 14.**
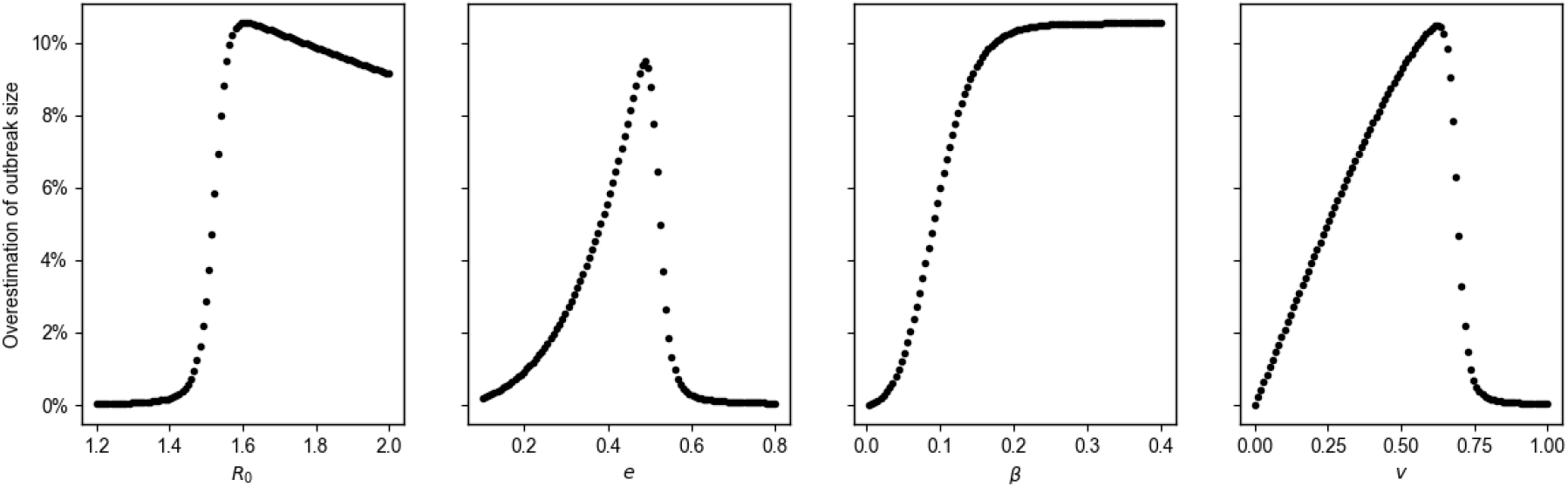
Overestimation of the outbreak size by employing a population-averaged vaccine SIR-model in contrast to a model that explicitly discriminates between vaccinated and unvaccinated individuals (see Eqs. (43)-(50)). Base parameters chosen here are vaccine efficacy against infection *e* = 0.5, recovery rate *β* = 1/7, reproduction rate ℛ_0_ = 1.55, initial fraction of vaccinated *v* = 2/3, initially infected *I*_0_ = 10^−4^. Population-averaged VE is given as *e* × *v*. Overestimations are shown for different ℛ_0_, VE *e*, recovery rate *β*, and fraction of vaccinated individuals *v* (from left to right).

**FIG. 15.**
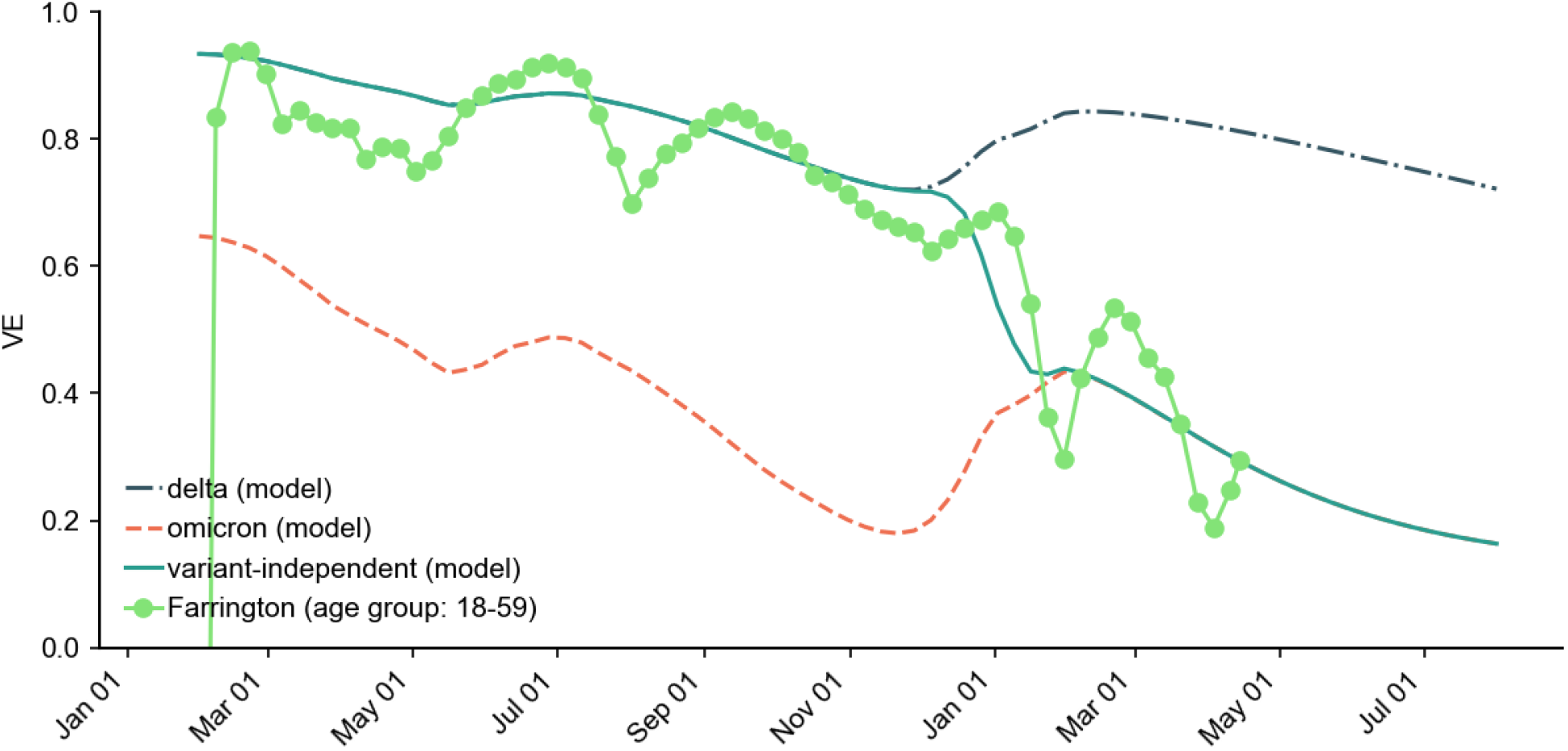
Retrospective comparison between vaccine efficacy obtained via Farrington’s method Eq. (51) from reported data and model scenario “high VE” and “medium reach” (cf. Fig. 2).

We first simulate (i) an *SI R* model with a population-averaged vaccine efficacy *v* × *e* as

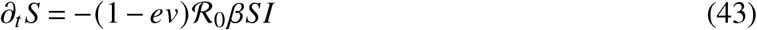

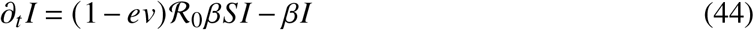

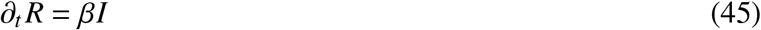

where *v* is the fraction of vaccinated individuals. This model is similar to Eqs. (1)-(4) in our main analysis, ignoring variants, contact modulations, and latency.

Second, we simulate (ii) an *S*-*S*_*V*_ -*I*-*I*_*V*_ -*R* model (subscript *V* marks vaccinated individuals) in which the vaccine efficacy affects only *S*_*V*_ individuals explicitly and breakthrough infections are counted separately, defined by

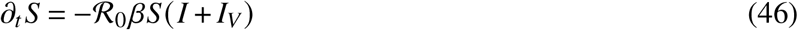

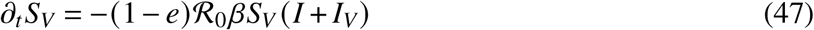

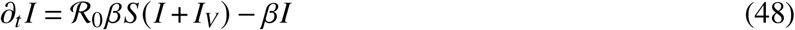

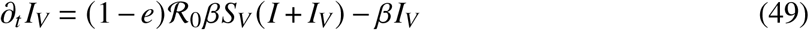

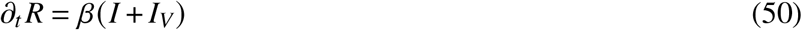

Note that in model (i), the fraction of vaccinated individuals is given by the parameter *v*, while in model (ii), the fraction of vaccinated individuals is implemented via initial conditions *S*(*t* = 0) = 1 − *v* − *I*_0_, *S*_*V*_ (*t* = 0) = *v*, and *I* (*t* = 0) = *I*_0_.

As can be seen in Fig. 14, the first model overestimates the outbreak size by approximately 10% in the worst case (with equal parameter values for both models), in a domain of parameter values that is in line with values chosen for our main analysis.

### 5.11. Retrospective evaluation of the population-wide vaccine efficacy

Using Farrington’s method [46], we retrospectively compute the per-calendar-week vaccine efficacy as

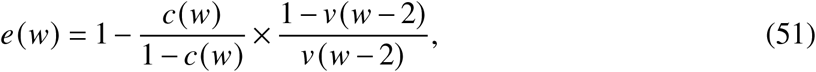

where we define as *c*(*w*) = Δ*C*_*V*_ (*w*)/(Δ*C*_*V*_ (*w*) + Δ*C*_*I*_ (*w*) the share of new infections in calendar week *w* and as *v* (*w*) the cumulative share of full vaccinations up to and including week *w* − 1 (note that in the equation above we shift *v*(*w*) by two weeks to account for the period until full immunity is reached). The share *v* of the vaccinated population is obtained from [21] and the share of breakthrough infections (not discriminating between boostered individuals and those that were fully vaccinated but not boostered) per calendar week from the German reporting system SurvStat [47, 48]. The data contains both symptomatic and asymptomatic infections. We find that the temporal evolution of vaccine efficacy as computed with Farrington’s method corresponds to our “high VE”, “medium reach” scenario (the latter being the assumption that about 80% of the individuals that received 2 doses in 2021 received a booster vaccination, as well), which is close to the empirically observed share of boostered individuals.

## Data Availability

All data produced in the present study are contained in the manuscript or available upon reasonable request to the authors.

## ACKNOWLEDGMENTS

We thank L.H. Wieler, L. Schaade, U. Buchholz, W. Haas, and C. Winklmayr for detailed discussions and substantial contributions that significantly influenced this work. B. F. M. is financially supported by the Joachim Herz Stiftung as an *Add-On Fellow for Interdisciplinary Life Science*.

---

**TABLE III.** Maximum possible relative risk (RR) of requiring intensive care for infections with Omicron vs. infections with Delta to keep ICU occupancy below a value of 4 800 beds.

| Inf.per. [d] | Omic. Lat. [d] | Booster Reach | Booster VE | Contact Reduction | max. RR |
| --- | --- | --- | --- | --- | --- |
| 2 | 1 | med. reach | low VE | None | 20%–25% |
| 2 | 1 | med. reach | low VE | -20%, Jan 31 to Mar 15 | 30%–35% |
| 2 | 1 | med. reach | low VE | -50%, Jan 31 to Feb 15 | 40%–45% |
| 2 | 1 | med. reach | low VE | -50%, Jan 31 to Feb 28 | 40%–45% |
| 2 | 1 | med. reach | low VE | -50%, Jan 31 to Mar 15 | 40%–45% |
| 2 | 1 | med. reach | low VE | -50%, Feb 15 to Mar 15 | 20%–25% |
| 2 | 1 | med. reach | high VE | None | 30%–35% |
| 2 | 1 | med. reach | high VE | -20%, Jan 31 to Mar 15 | 50%–55% |
| 2 | 1 | med. reach | high VE | -50%, Jan 31 to Feb 15 | 55%–60% |
| 2 | 1 | med. reach | high VE | -50%, Jan 31 to Feb 28 | 55%–60% |
| 2 | 1 | med. reach | high VE | -50%, Jan 31 to Mar 15 | 55%–60% |
| 2 | 1 | med. reach | high VE | -50%, Feb 15 to Mar 15 | 35%–40% |
| 2 | 1 | high reach | low VE | None | 20%–25% |
| 2 | 1 | high reach | low VE | -20%, Jan 31 to Mar 15 | 30%–35% |
| 2 | 1 | high reach | low VE | -50%, Jan 31 to Feb 15 | 40%–45% |
| 2 | 1 | high reach | low VE | -50%, Jan 31 to Feb 28 | 40%–45% |
| 2 | 1 | high reach | low VE | -50%, Jan 31 to Mar 15 | 40%–45% |
| 2 | 1 | high reach | low VE | -50%, Feb 15 to Mar 15 | 20%–25% |
| 2 | 1 | high reach | high VE | None | 35%–40% |
| 2 | 1 | high reach | high VE | -20%, Jan 31 to Mar 15 | 50%–55% |
| 2 | 1 | high reach | high VE | -50%, Jan 31 to Feb 15 | 55%–60% |
| 2 | 1 | high reach | high VE | -50%, Jan 31 to Feb 28 | 55%–60% |
| 2 | 1 | high reach | high VE | -50%, Jan 31 to Mar 15 | 55%–60% |
| 2 | 1 | high reach | high VE | -50%, Feb 15 to Mar 15 | 40%–45% |

**TABLE IV.** Maximum possible relative risk (RR) of requiring intensive care for infections with Omicron vs. infections with Delta to keep ICU occupancy below a value of 4 800 beds.

| Inf.per. [d] | Omic. Lat. [d] | Booster Reach | Booster VE | Cotact Reduction | max. RR |
| --- | --- | --- | --- | --- | --- |
| 2 | 2 | med. reach | low VE | None | 15%–20% |
| 2 | 2 | med. reach | low VE | -20%, Jan 31 to Mar 15 | 20%–25% |
| 2 | 2 | med. reach | low VE | -50%, Jan 31 to Feb 15 | 30%–35% |
| 2 | 2 | med. reach | low VE | -50%, Jan 31 to Feb 28 | 30%–35% |
| 2 | 2 | med. reach | low VE | -50%, Jan 31 to Mar 15 | 30%–35% |
| 2 | 2 | med. reach | low VE | -50%, Feb 15 to Mar 15 | 15%–20% |
| 2 | 2 | med. reach | high VE | None | 15%–20% |
| 2 | 2 | med. reach | high VE | -20%, Jan 31 to Mar 15 | 30%–35% |
| 2 | 2 | med. reach | high VE | -50%, Jan 31 to Feb 15 | 40%–45% |
| 2 | 2 | med. reach | high VE | -50%, Jan 31 to Feb 28 | 40%–45% |
| 2 | 2 | med. reach | high VE | -50%, Jan 31 to Mar 15 | 40%–45% |
| 2 | 2 | med. reach | high VE | -50%, Feb 15 to Mar 15 | 20%–25% |
| 2 | 2 | high reach | low VE | None | 15%–20% |
| 2 | 2 | high reach | low VE | -20%, Jan 31 to Mar 15 | 20%–25% |
| 2 | 2 | high reach | low VE | -50%, Jan 31 to Feb 15 | 30%–35% |
| 2 | 2 | high reach | low VE | -50%, Jan 31 to Feb 28 | 30%–35% |
| 2 | 2 | high reach | low VE | -50%, Jan 31 to Mar 15 | 30%–35% |
| 2 | 2 | high reach | low VE | -50%, Feb 15 to Mar 15 | 15%–20% |
| 2 | 2 | high reach | high VE | None | 20%–25% |
| 2 | 2 | high reach | high VE | -20%, Jan 31 to Mar 15 | 30%–35% |
| 2 | 2 | high reach | high VE | -50%, Jan 31 to Feb 15 | 40%–45% |
| 2 | 2 | high reach | high VE | -50%, Jan 31 to Feb 28 | 40%–45% |
| 2 | 2 | high reach | high VE | -50%, Jan 31 to Mar 15 | 40%–45% |
| 2 | 2 | high reach | high VE | -50%, Feb 15 to Mar 15 | 20%–25% |

**TABLE V.** Maximum possible relative risk (RR) of requiring intensive care for infections with Omicron vs. infections with Delta to keep ICU occupancy below a value of 4 800 beds.

| Inf.per. [d] | Omic. Lat. [d] | Booster Reach | Booster VE | Contact Reduction | max. RR |
| --- | --- | --- | --- | --- | --- |
| 3 | 1 | med. reach | low VE | None | 15%–20% |
| 3 | 1 | med. reach | low VE | -20%, Jan 31 to Mar 15 | 25%–30% |
| 3 | 1 | med. reach | low VE | -50%, Jan 31 to Feb 15 | 35%–40% |
| 3 | 1 | med. reach | low VE | -50%, Jan 31 to Feb 28 | 35%–40% |
| 3 | 1 | med. reach | low VE | -50%, Jan 31 to Mar 15 | 35%–40% |
| 3 | 1 | med. reach | low VE | -50%, Feb 15 to Mar 15 | 15%–20% |
| 3 | 1 | med. reach | high VE | None | 20%–25% |
| 3 | 1 | med. reach | high VE | -20%, Jan 31 to Mar 15 | 40%–45% |
| 3 | 1 | med. reach | high VE | -50%, Jan 31 to Feb 15 | 45%–50% |
| 3 | 1 | med. reach | high VE | -50%, Jan 31 to Feb 28 | 45%–50% |
| 3 | 1 | med. reach | high VE | -50%, Jan 31 to Mar 15 | 45%–50% |
| 3 | 1 | med. reach | high VE | -50%, Feb 15 to Mar 15 | 25%–30% |
| 3 | 1 | high reach | low VE | None | 15%–20% |
| 3 | 1 | high reach | low VE | -20%, Jan 31 to Mar 15 | 25%–30% |
| 3 | 1 | high reach | low VE | -50%, Jan 31 to Feb 15 | 35%–40% |
| 3 | 1 | high reach | low VE | -50%, Jan 31 to Feb 28 | 35%–40% |
| 3 | 1 | high reach | low VE | -50%, Jan 31 to Mar 15 | 35%–40% |
| 3 | 1 | high reach | low VE | -50%, Feb 15 to Mar 15 | 20%–25% |
| 3 | 1 | high reach | high VE | None | 25%–30% |
| 3 | 1 | high reach | high VE | -20%, Jan 31 to Mar 15 | 40%–45% |
| 3 | 1 | high reach | high VE | -50%, Jan 31 to Feb 15 | 45%–50% |
| 3 | 1 | high reach | high VE | -50%, Jan 31 to Feb 28 | 45%–50% |
| 3 | 1 | high reach | high VE | -50%, Jan 31 to Mar 15 | 45%–50% |
| 3 | 1 | high reach | high VE | -50%, Feb 15 to Mar 15 | 30%–35% |

**TABLE VI.** Maximum possible relative risk (RR) of requiring intensive care for infections with Omicron vs. infections with Delta to keep ICU occupancy below a value of 4 800 beds.

| Inf.per. [d] | Omic. Lat. [d] | Booster Reach | Booster VE | Contact Reduction | max. RR |
| --- | --- | --- | --- | --- | --- |
| 3 | 2 | med. reach | low VE | None | 10%–15% |
| 3 | 2 | med. reach | low VE | -20%, Jan 31 to Mar 15 | 15%–20% |
| 3 | 2 | med. reach | low VE | -50%, Jan 31 to Feb 15 | 20%–25% |
| 3 | 2 | med. reach | low VE | -50%, Jan 31 to Feb 28 | 25%–30% |
| 3 | 2 | med. reach | low VE | -50%, Jan 31 to Mar 15 | 25%–30% |
| 3 | 2 | med. reach | low VE | -50%, Feb 15 to Mar 15 | 15%–20% |
| 3 | 2 | med. reach | high VE | None | 15%–20% |
| 3 | 2 | med. reach | high VE | -20%, Jan 31 to Mar 15 | 25%–30% |
| 3 | 2 | med. reach | high VE | -50%, Jan 31 to Feb 15 | 35%–40% |
| 3 | 2 | med. reach | high VE | -50%, Jan 31 to Feb 28 | 35%–40% |
| 3 | 2 | med. reach | high VE | -50%, Jan 31 to Mar 15 | 35%–40% |
| 3 | 2 | med. reach | high VE | -50%, Feb 15 to Mar 15 | 15%–20% |
| 3 | 2 | high reach | low VE | None | 10%–15% |
| 3 | 2 | high reach | low VE | -20%, Jan 31 to Mar 15 | 15%–20% |
| 3 | 2 | high reach | low VE | -50%, Jan 31 to Feb 15 | 25%–30% |
| 3 | 2 | high reach | low VE | -50%, Jan 31 to Feb 28 | 25%–30% |
| 3 | 2 | high reach | low VE | -50%, Jan 31 to Mar 15 | 25%–30% |
| 3 | 2 | high reach | low VE | -50%, Feb 15 to Mar 15 | 15%–20% |
| 3 | 2 | high reach | high VE | None | 15%–20% |
| 3 | 2 | high reach | high VE | -20%, Jan 31 to Mar 15 | 25%–30% |
| 3 | 2 | high reach | high VE | -50%, Jan 31 to Feb 15 | 35%–40% |
| 3 | 2 | high reach | high VE | -50%, Jan 31 to Feb 28 | 35%–40% |
| 3 | 2 | high reach | high VE | -50%, Jan 31 to Mar 15 | 35%–40% |
| 3 | 2 | high reach | high VE | -50%, Feb 15 to Mar 15 | 20%–25% |

**TABLE VII.** Outbreak sizes expected according to model by April 1, 2022 (cumulative reported cases of Omicron infections assuming constant unreported cases).

| Inf.per. [d] | Omic. Lat. [d] | Booster Reach | Booster VE | Contact Reduction | Om. Cases Total [Mil] |
| --- | --- | --- | --- | --- | --- |
| 2 | 1 | med. reach | low VE | None | 18.9 |
| 2 | 1 | med. reach | low VE | -20%, Jan 31 to Mar 15 | 11.5 |
| 2 | 1 | med. reach | low VE | -50%, Jan 31 to Feb 15 | 10.2 |
| 2 | 1 | med. reach | low VE | -50%, Jan 31 to Feb 28 | 6.6 |
| 2 | 1 | med. reach | low VE | -50%, Jan 31 to Mar 15 | 6.3 |
| 2 | 1 | med. reach | low VE | -50%, Feb 15 to Mar 15 | 12.9 |
| 2 | 1 | med. reach | high VE | None | 11.5 |
| 2 | 1 | med. reach | high VE | -20%, Jan 31 to Mar 15 | 5.7 |
| 2 | 1 | med. reach | high VE | -50%, Jan 31 to Feb 15 | 4.6 |
| 2 | 1 | med. reach | high VE | -50%, Jan 31 to Feb 28 | 3.8 |
| 2 | 1 | med. reach | high VE | -50%, Jan 31 to Mar 15 | 3.7 |
| 2 | 1 | med. reach | high VE | -50%, Feb 15 to Mar 15 | 6.6 |
| 2 | 1 | high reach | low VE | None | 17.4 |
| 2 | 1 | high reach | low VE | -20%, Jan 31 to Mar 15 | 10.5 |
| 2 | 1 | high reach | low VE | -50%, Jan 31 to Feb 15 | 8.8 |
| 2 | 1 | high reach | low VE | -50%, Jan 31 to Feb 28 | 6.4 |
| 2 | 1 | high reach | low VE | -50%, Jan 31 to Mar 15 | 6.2 |
| 2 | 1 | high reach | low VE | -50%, Feb 15 to Mar 15 | 12.1 |
| 2 | 1 | high reach | high VE | None | 9.4 |
| 2 | 1 | high reach | high VE | -20%, Jan 31 to Mar 15 | 5.2 |
| 2 | 1 | high reach | high VE | -50%, Jan 31 to Feb 15 | 4.1 |
| 2 | 1 | high reach | high VE | -50%, Jan 31 to Feb 28 | 3.7 |
| 2 | 1 | high reach | high VE | -50%, Jan 31 to Mar 15 | 3.7 |
| 2 | 1 | high reach | high VE | -50%, Feb 15 to Mar 15 | 6.2 |

**TABLE VIII.** Outbreak sizes expected according to model by April 1, 2022 (cumulative reported cases of Omicron infections assuming constant unreported cases).

| Inf.per. [d] | Omic. Lat. [d] | Booster Reach | Booster VE | Contact Reduction | Om. Cases Total [Mil] |
| --- | --- | --- | --- | --- | --- |
| 2 | 2 | med. reach | low VE | None | 23.6 |
| 2 | 2 | med. reach | low VE | -20%, Jan 31 to Mar 15 | 16.9 |
| 2 | 2 | med. reach | low VE | -50%, Jan 31 to Feb 15 | 16.0 |
| 2 | 2 | med. reach | low VE | -50%, Jan 31 to Feb 28 | 10.1 |
| 2 | 2 | med. reach | low VE | -50%, Jan 31 to Mar 15 | 8.9 |
| 2 | 2 | med. reach | low VE | -50%, Feb 15 to Mar 15 | 17.5 |
| 2 | 2 | med. reach | high VE | None | 17.5 |
| 2 | 2 | med. reach | high VE | -20%, Jan 31 to Mar 15 | 10.4 |
| 2 | 2 | med. reach | high VE | -50%, Jan 31 to Feb 15 | 8.8 |
| 2 | 2 | med. reach | high VE | -50%, Jan 31 to Feb 28 | 5.9 |
| 2 | 2 | med. reach | high VE | -50%, Jan 31 to Mar 15 | 5.5 |
| 2 | 2 | med. reach | high VE | -50%, Feb 15 to Mar 15 | 10.7 |
| 2 | 2 | high reach | low VE | None | 22.4 |
| 2 | 2 | high reach | low VE | -20%, Jan 31 to Mar 15 | 15.7 |
| 2 | 2 | high reach | low VE | -50%, Jan 31 to Feb 15 | 14.3 |
| 2 | 2 | high reach | low VE | -50%, Jan 31 to Feb 28 | 9.5 |
| 2 | 2 | high reach | low VE | -50%, Jan 31 to Mar 15 | 8.7 |
| 2 | 2 | high reach | low VE | -50%, Feb 15 to Mar 15 | 16.6 |
| 2 | 2 | high reach | high VE | None | 15.4 |
| 2 | 2 | high reach | high VE | -20%, Jan 31 to Mar 15 | 9.2 |
| 2 | 2 | high reach | high VE | -50%, Jan 31 to Feb 15 | 7.4 |
| 2 | 2 | high reach | high VE | -50%, Jan 31 to Feb 28 | 5.6 |
| 2 | 2 | high reach | high VE | -50%, Jan 31 to Mar 15 | 5.4 |
| 2 | 2 | high reach | high VE | -50%, Feb 15 to Mar 15 | 10.0 |

**TABLE IX.** Outbreak sizes expected according to model by April 1, 2022 (cumulative reported cases of Omicron infections assuming constant unreported cases).

| Inf.per. [d] | Omic. Lat. [d] | Booster Reach | Booster VE | Contact Reduction | Om. Cases Total [Mil] |
| --- | --- | --- | --- | --- | --- |
| 3 | 1 | med. reach | low VE | None | 22.6 |
| 3 | 1 | med. reach | low VE | -20%, Jan 31 to Mar 15 | 15.2 |
| 3 | 1 | med. reach | low VE | -50%, Jan 31 to Feb 15 | 14.9 |
| 3 | 1 | med. reach | low VE | -50%, Jan 31 to Feb 28 | 8.7 |
| 3 | 1 | med. reach | low VE | -50%, Jan 31 to Mar 15 | 7.5 |
| 3 | 1 | med. reach | low VE | -50%, Feb 15 to Mar 15 | 15.4 |
| 3 | 1 | med. reach | high VE | None | 15.8 |
| 3 | 1 | med. reach | high VE | -20%, Jan 31 to Mar 15 | 8.6 |
| 3 | 1 | med. reach | high VE | -50%, Jan 31 to Feb 15 | 7.5 |
| 3 | 1 | med. reach | high VE | -50%, Jan 31 to Feb 28 | 4.9 |
| 3 | 1 | med. reach | high VE | -50%, Jan 31 to Mar 15 | 4.6 |
| 3 | 1 | med. reach | high VE | -50%, Feb 15 to Mar 15 | 8.9 |
| 3 | 1 | high reach | low VE | None | 21.2 |
| 3 | 1 | high reach | low VE | -20%, Jan 31 to Mar 15 | 14.0 |
| 3 | 1 | high reach | low VE | -50%, Jan 31 to Feb 15 | 13.1 |
| 3 | 1 | high reach | low VE | -50%, Jan 31 to Feb 28 | 8.1 |
| 3 | 1 | high reach | low VE | -50%, Jan 31 to Mar 15 | 7.3 |
| 3 | 1 | high reach | low VE | -50%, Feb 15 to Mar 15 | 14.5 |
| 3 | 1 | high reach | high VE | None | 13.5 |
| 3 | 1 | high reach | high VE | -20%, Jan 31 to Mar 15 | 7.6 |
| 3 | 1 | high reach | high VE | -50%, Jan 31 to Feb 15 | 6.2 |
| 3 | 1 | high reach | high VE | -50%, Jan 31 to Feb 28 | 4.7 |
| 3 | 1 | high reach | high VE | -50%, Jan 31 to Mar 15 | 4.5 |
| 3 | 1 | high reach | high VE | -50%, Feb 15 to Mar 15 | 8.2 |

**TABLE X.** Outbreak sizes expected according to model by April 1, 2022 (cumulative reported cases of Omicron infections assuming constant unreported cases).

| Inf.per. [d] | Omic. Lat. [d] | Booster Reach | Booster VE | Contact Reduction | Om. Cases Total [Mil] |
| --- | --- | --- | --- | --- | --- |
| 3 | 2 | med. reach | low VE | None | 26.5 |
| 3 | 2 | med. reach | low VE | -20%, Jan 31 to Mar 15 | 20.3 |
| 3 | 2 | med. reach | low VE | -50%, Jan 31 to Feb 15 | 20.1 |
| 3 | 2 | med. reach | low VE | -50%, Jan 31 to Feb 28 | 13.0 |
| 3 | 2 | med. reach | low VE | -50%, Jan 31 to Mar 15 | 10.6 |
| 3 | 2 | med. reach | low VE | -50%, Feb 15 to Mar 15 | 19.8 |
| 3 | 2 | med. reach | high VE | None | 21.1 |
| 3 | 2 | med. reach | high VE | -20%, Jan 31 to Mar 15 | 13.8 |
| 3 | 2 | med. reach | high VE | -50%, Jan 31 to Feb 15 | 12.7 |
| 3 | 2 | med. reach | high VE | -50%, Jan 31 to Feb 28 | 7.9 |
| 3 | 2 | med. reach | high VE | -50%, Jan 31 to Mar 15 | 6.8 |
| 3 | 2 | med. reach | high VE | -50%, Feb 15 to Mar 15 | 13.2 |
| 3 | 2 | high reach | low VE | None | 25.4 |
| 3 | 2 | high reach | low VE | -20%, Jan 31 to Mar 15 | 19.1 |
| 3 | 2 | high reach | low VE | -50%, Jan 31 to Feb 15 | 18.4 |
| 3 | 2 | high reach | low VE | -50%, Jan 31 to Feb 28 | 12.1 |
| 3 | 2 | high reach | low VE | -50%, Jan 31 to Mar 15 | 10.2 |
| 3 | 2 | high reach | low VE | -50%, Feb 15 to Mar 15 | 18.9 |
| 3 | 2 | high reach | high VE | None | 19.0 |
| 3 | 2 | high reach | high VE | -20%, Jan 31 to Mar 15 | 12.3 |
| 3 | 2 | high reach | high VE | -50%, Jan 31 to Feb 15 | 10.7 |
| 3 | 2 | high reach | high VE | -50%, Jan 31 to Feb 28 | 7.2 |
| 3 | 2 | high reach | high VE | -50%, Jan 31 to Mar 15 | 6.5 |
| 3 | 2 | high reach | high VE | -50%, Feb 15 to Mar 15 | 12.2 |

**TABLE XI.** Maximum values of all local maxima of incidence, hospitalization incidence and ITS occupancy in the period January 2022 to April 2022. Curves with entries “−” show no local maxima. Here, RR Hosp. = 0.35 and RR ICU = 0.15 were assumed.

| $\beta^{-1}$ [d] | $\omega_o^{-1}$ [d] | Reach | Booster VE | Cont.red. | $\max(J_C)/10^3$ | $\max(J_H)/10^3$ | $\max(U)/10^3$ |
| --- | --- | --- | --- | --- | --- | --- | --- |
| 2 | 1 | med. | low VE | None | 437 | 2.6 | 3.7 |
| 2 | 1 | med. | low VE | -20%, 01.31.–03.15. | 268 | 1.7 | 2.5 |
| 2 | 1 | med. | low VE | -50%, 01.31.–02.15. | 233 | 1.5 | 2.3 |
| 2 | 1 | med. | low VE | -50%, 01.31.–02.28. | 233 | 1.5 | 2.3 |
| 2 | 1 | med. | low VE | -50%, 01.31.–03.15. | 233 | 1.5 | 2.3 |
| 2 | 1 | med. | low VE | -50%, 02.15.–03.15. | 416 | 2.5 | 3.3 |
| 2 | 1 | med. | high VE | None | 198 | 1.5 | 2.3 |
| 2 | 1 | med. | high VE | -20%, 01.31.–03.15. | 135 | 1.1 | – |
| 2 | 1 | med. | high VE | -50%, 01.31.–02.15. | 127 | 1.0 | – |
| 2 | 1 | med. | high VE | -50%, 01.31.–02.28. | 127 | 1.0 | – |
| 2 | 1 | med. | high VE | -50%, 01.31.–03.15. | 127 | 1.0 | – |
| 2 | 1 | med. | high VE | -50%, 02.15.–03.15. | 183 | 1.4 | 2.1 |
| 2 | 1 | high | low VE | None | 392 | 2.4 | 3.4 |
| 2 | 1 | high | low VE | -20%, 01.31.–03.15. | 260 | 1.6 | 2.5 |
| 2 | 1 | high | low VE | -50%, 01.31.–02.15. | 231 | 1.5 | 2.3 |
| 2 | 1 | high | low VE | -50%, 01.31.–02.28. | 231 | 1.5 | 2.3 |
| 2 | 1 | high | low VE | -50%, 01.31.–03.15. | 231 | 1.5 | 2.3 |
| 2 | 1 | high | low VE | -50%, 02.15.–03.15. | 382 | 2.3 | 3.1 |
| 2 | 1 | high | high VE | None | 166 | 1.3 | 2.1 |
| 2 | 1 | high | high VE | -20%, 01.31.–03.15. | 132 | 1.1 | – |
| 2 | 1 | high | high VE | -50%, 01.31.–02.15. | 126 | 1.0 | – |
| 2 | 1 | high | high VE | -50%, 01.31.–02.28. | 126 | 1.0 | – |
| 2 | 1 | high | high VE | -50%, 01.31.–03.15. | 126 | 1.0 | – |
| 2 | 1 | high | high VE | -50%, 02.15.–03.15. | 165 | 1.3 | 2.1 |

**TABLE XII.**
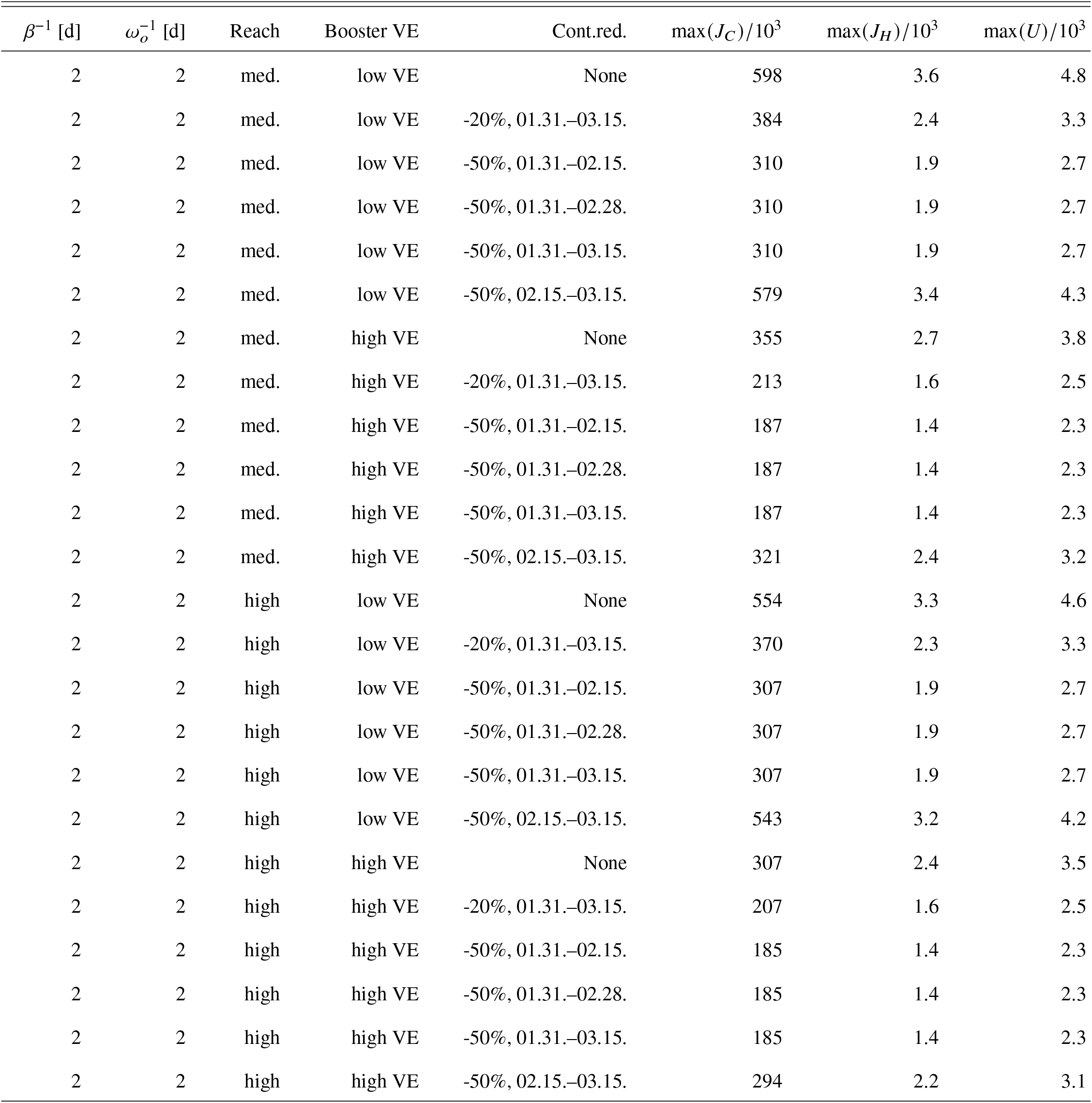
Maximum values of all local maxima of incidence, hospitalization incidence and ITS occupancy in the period January 2022 to April 2022. Curves with entries “−” show no local maxima. Here, RR Hosp. = 0.35 and RR ICU = 0.15 were assumed.

| $\beta^{-1}$ [d] | $\omega_o^{-1}$ [d] | Reach | Booster VE | Cont.red. | $\max(J_C)/10^3$ | $\max(J_H)/10^3$ | $\max(U)/10^3$ |
| --- | --- | --- | --- | --- | --- | --- | --- |
| 2 | 2 | med. | low VE | None | 598 | 3.6 | 4.8 |
| 2 | 2 | med. | low VE | -20%, 01.31.–03.15. | 384 | 2.4 | 3.3 |
| 2 | 2 | med. | low VE | -50%, 01.31.–02.15. | 310 | 1.9 | 2.7 |
| 2 | 2 | med. | low VE | -50%, 01.31.–02.28. | 310 | 1.9 | 2.7 |
| 2 | 2 | med. | low VE | -50%, 01.31.–03.15. | 310 | 1.9 | 2.7 |
| 2 | 2 | med. | low VE | -50%, 02.15.–03.15. | 579 | 3.4 | 4.3 |
| 2 | 2 | med. | high VE | None | 355 | 2.7 | 3.8 |
| 2 | 2 | med. | high VE | -20%, 01.31.–03.15. | 213 | 1.6 | 2.5 |
| 2 | 2 | med. | high VE | -50%, 01.31.–02.15. | 187 | 1.4 | 2.3 |
| 2 | 2 | med. | high VE | -50%, 01.31.–02.28. | 187 | 1.4 | 2.3 |
| 2 | 2 | med. | high VE | -50%, 01.31.–03.15. | 187 | 1.4 | 2.3 |
| 2 | 2 | med. | high VE | -50%, 02.15.–03.15. | 321 | 2.4 | 3.2 |
| 2 | 2 | high | low VE | None | 554 | 3.3 | 4.6 |
| 2 | 2 | high | low VE | -20%, 01.31.–03.15. | 370 | 2.3 | 3.3 |
| 2 | 2 | high | low VE | -50%, 01.31.–02.15. | 307 | 1.9 | 2.7 |
| 2 | 2 | high | low VE | -50%, 01.31.–02.28. | 307 | 1.9 | 2.7 |
| 2 | 2 | high | low VE | -50%, 01.31.–03.15. | 307 | 1.9 | 2.7 |
| 2 | 2 | high | low VE | -50%, 02.15.–03.15. | 543 | 3.2 | 4.2 |
| 2 | 2 | high | high VE | None | 307 | 2.4 | 3.5 |
| 2 | 2 | high | high VE | -20%, 01.31.–03.15. | 207 | 1.6 | 2.5 |
| 2 | 2 | high | high VE | -50%, 01.31.–02.15. | 185 | 1.4 | 2.3 |
| 2 | 2 | high | high VE | -50%, 01.31.–02.28. | 185 | 1.4 | 2.3 |
| 2 | 2 | high | high VE | -50%, 01.31.–03.15. | 185 | 1.4 | 2.3 |
| 2 | 2 | high | high VE | -50%, 02.15.–03.15. | 294 | 2.2 | 3.1 |

**TABLE XIII.** Maximum values of all local maxima of incidence, hospitalization incidence and ITS occupancy in the period January 2022 to April 2022. Curves with entries “−” show no local maxima. Here, RR Hosp. = 0.35 and RR ICU = 0.15 were assumed.

| $\beta^{-1}$ [d] | $\omega_o^{-1}$ [d] | Reach | Booster VE | Cont.red. | $\max(J_C)/10^3$ | $\max(J_H)/10^3$ | $\max(U)/10^3$ |
| --- | --- | --- | --- | --- | --- | --- | --- |
| 3 | 1 | med. | low VE | None | 539 | 3.2 | 4.4 |
| 3 | 1 | med. | low VE | -20%, 01.31.–03.15. | 312 | 1.9 | 2.8 |
| 3 | 1 | med. | low VE | -50%, 01.31.–02.15. | 343 | 1.9 | 2.4 |
| 3 | 1 | med. | low VE | -50%, 01.31.–02.28. | 252 | 1.6 | 2.4 |
| 3 | 1 | med. | low VE | -50%, 01.31.–03.15. | 252 | 1.6 | 2.4 |
| 3 | 1 | med. | low VE | -50%, 02.15.–03.15. | 498 | 2.9 | 3.7 |
| 3 | 1 | med. | high VE | None | 302 | 2.3 | 3.3 |
| 3 | 1 | med. | high VE | -20%, 01.31.–03.15. | 166 | 1.3 | 2.2 |
| 3 | 1 | med. | high VE | -50%, 01.31.–02.15. | 150 | 1.2 | – |
| 3 | 1 | med. | high VE | -50%, 01.31.–02.28. | 150 | 1.2 | – |
| 3 | 1 | med. | high VE | -50%, 01.31.–03.15. | 150 | 1.2 | – |
| 3 | 1 | med. | high VE | -50%, 02.15.–03.15. | 254 | 1.9 | 2.6 |
| 3 | 1 | high | low VE | None | 490 | 2.9 | 4.2 |
| 3 | 1 | high | low VE | -20%, 01.31.–03.15. | 299 | 1.9 | 2.8 |
| 3 | 1 | high | low VE | -50%, 01.31.–02.15. | 249 | 1.6 | 2.4 |
| 3 | 1 | high | low VE | -50%, 01.31.–02.28. | 249 | 1.6 | 2.4 |
| 3 | 1 | high | low VE | -50%, 01.31.–03.15. | 249 | 1.6 | 2.4 |
| 3 | 1 | high | low VE | -50%, 02.15.–03.15. | 462 | 2.7 | 3.6 |
| 3 | 1 | high | high VE | None | 249 | 1.9 | 2.9 |
| 3 | 1 | high | high VE | -20%, 01.31.–03.15. | 162 | 1.3 | – |
| 3 | 1 | high | high VE | -50%, 01.31.–02.15. | 148 | 1.2 | – |
| 3 | 1 | high | high VE | -50%, 01.31.–02.28. | 148 | 1.2 | – |
| 3 | 1 | high | high VE | -50%, 01.31.–03.15. | 148 | 1.2 | – |
| 3 | 1 | high | high VE | -50%, 02.15.–03.15. | 231 | 1.8 | 2.5 |

**TABLE XIV.** Maximum values of all local maxima of incidence, hospitalization incidence and ITS occupancy in the period January 2022 to April 2022. Curves with entries “−” show no local maxima. Here, RR Hosp. = 0.35 and RR ICU = 0.15 were assumed.

| $\beta^{-1}$ [d] | $\omega_o^{-1}$ [d] | Reach | Booster VE | Cont.red. | $\max(J_C)/10^3$ | $\max(J_H)/10^3$ | $\max(U)/10^3$ |
| --- | --- | --- | --- | --- | --- | --- | --- |
| 3 | 2 | med. | low VE | None | 680 | 4.0 | 5.3 |
| 3 | 2 | med. | low VE | -20%, 01.31.–03.15. | 438 | 2.7 | 3.8 |
| 3 | 2 | med. | low VE | -50%, 01.31.–02.15. | 385 | 2.2 | 3.1 |
| 3 | 2 | med. | low VE | -50%, 01.31.–02.28. | 325 | 2.0 | 2.8 |
| 3 | 2 | med. | low VE | -50%, 01.31.–03.15. | 325 | 2.0 | 2.8 |
| 3 | 2 | med. | low VE | -50%, 02.15.–03.15. | 647 | 3.8 | 4.7 |
| 3 | 2 | med. | high VE | None | 450 | 3.4 | 4.7 |
| 3 | 2 | med. | high VE | -20%, 01.31.–03.15. | 254 | 2.0 | 2.9 |
| 3 | 2 | med. | high VE | -50%, 01.31.–02.15. | 208 | 1.6 | 2.4 |
| 3 | 2 | med. | high VE | -50%, 01.31.–02.28. | 208 | 1.6 | 2.4 |
| 3 | 2 | med. | high VE | -50%, 01.31.–03.15. | 208 | 1.6 | 2.4 |
| 3 | 2 | med. | high VE | -50%, 02.15.–03.15. | 391 | 2.9 | 3.7 |
| 3 | 2 | high | low VE | None | 635 | 3.8 | 5.2 |
| 3 | 2 | high | low VE | -20%, 01.31.–03.15. | 416 | 2.6 | 3.7 |
| 3 | 2 | high | low VE | -50%, 01.31.–02.15. | 332 | 2.0 | 2.9 |
| 3 | 2 | high | low VE | -50%, 01.31.–02.28. | 321 | 2.0 | 2.8 |
| 3 | 2 | high | low VE | -50%, 01.31.–03.15. | 321 | 2.0 | 2.8 |
| 3 | 2 | high | low VE | -50%, 02.15.–03.15. | 610 | 3.6 | 4.6 |
| 3 | 2 | high | high VE | None | 394 | 3.0 | 4.4 |
| 3 | 2 | high | high VE | -20%, 01.31.–03.15. | 243 | 1.9 | 2.8 |
| 3 | 2 | high | high VE | -50%, 01.31.–02.15. | 206 | 1.6 | 2.4 |
| 3 | 2 | high | high VE | -50%, 01.31.–02.28. | 206 | 1.6 | 2.4 |
| 3 | 2 | high | high VE | -50%, 01.31.–03.15. | 206 | 1.6 | 2.4 |
| 3 | 2 | high | high VE | -50%, 02.15.–03.15. | 360 | 2.7 | 3.6 |

